# Efficacy and safety of semaglutide for obesity and hyperphagia in adults with Prader-Willi syndrome

**DOI:** 10.64898/2026.06.11.26354795

**Authors:** Shahd Ahmed, Nicola Bridges, Anthony P. Goldstone

## Abstract

**Context:** Prader-Willi syndrome is a genetic neurodevelopmental disorder characterized by hyperphagia and early-onset obesity from hypothalamic dysfunction with endocrinopathies and learning disability. Management is challenging with strict control of the food environment needed. While newer glucagon-like peptide-1 receptor agonists, such as semaglutide, have efficacy in non-PWS obesity, there have been limited case reports in PWS.

**Objective/Design/Setting:** Retrospective, observational cohort of 12 adults with PWS and overweight/obesity treated with semaglutide at a UK academic hospital centre specialist clinic.

**Patients:** Mean ± SD age 28.3 ± 10.1 years, 83% female, BMI 46.6 ± 8.2kg/m², 75% type 2 diabetes mellitus.

**Intervention:** Median follow-up 17.2 months (range 8.7-36.1) with median semaglutide dose 2.4mg once weekly (1.0-2.4).

**Results:** There was no significant overall weight loss on semaglutide, but there was stabilisation of the weight gain prior to treatment over previous 12.4 months (7.6-23.0): post -3.1 ± 9.9% vs. pre +5.7 ± 5.6%: d -0.72, P=0.037. There was a significant decrease in hyperphagia on semaglutide from Hyperphagia Questionnaire for Clinical Trials (n=11, -7.3 ± 6.1 (max 36), d -1.19, P=0.003), having been stable before treatment. HbA1c improved in those with elevated baseline levels (n=6, -4.2 ± 4.9%, d -0.74, P=0.13). Mild gastrointestinal side effects were seen in 25% but did not lead to discontinuation.

**Conclusions:** In this small open-label, observational cohort of adults with PWS, semaglutide produced weight maintenance though not weight loss, but appeared to reduce hyperphagia, and improved glycaemic control, with good tolerability. Larger placebo-controlled trials are needed to confirm these findings in adults and adolescents with PWS, especially in those without T2DM, where efficacy may be greater.

## 1. INTRODUCTION

### 1.1. Prader-Willi Syndrome

Prader Willi syndrome (PWS) is a rare orphan disease and complex genetic neurodevelopmental disorder, with no licensed treatment for the primary unmet need of controlling hyperphagia and obesity. Nutritional and growth phases in children and adolescents with PWS are well described, with weight gain and hyperphagia leading to obesity later on in childhood if access to food is uncontrolled, with premature mortality primarily due to obesity/diabetes-related complications in adulthood, including cardiorespiratory disease, chest infections, and even choking and gastric rupture related to acute overeating episodes (1–6).

PWS has a birth incidence of approximately 1 in 10,000-30,000 that results from loss of paternal expression of several maternally imprinted genes on chromosome 15q11-13, most commonly due to a paternal deletion or maternal uniparental disomy, with rarer causes such as imprinting defects, translocations and microdeletions (7, 8). The resulting lack of expression of paternal genes within this region disrupts hypothalamic and more widespread neural development and function (9–13).

Hypotonia, poor feeding and growth are characteristic features in the neonatal period, followed by reduced calorie requirement and subsequent hyperphagia and risk of rapid weight gain later in childhood (2, 11, 14). Children and adolescents with PWS usually display developmental delay and mild learning disability, and can develop endocrinopathies (especially growth hormone deficiency and hypogonadism), scoliosis, respiratory difficulties (both central and obstructive sleep apnoea), challenging behaviours (especially temper tantrums around access to food), skin picking, and mental health issues especially into adulthood (1, 6, 15, 16).

### 1.2. Hyperphagia and Obesity in PWS

Hyperphagia in PWS can manifest as increased hunger, impaired satiety, preoccupation with food, bargaining or manipulating to get more food at meals, foraging through trash for food, eating rotten food or non-food items, getting up at night to food seek, being persistent in asking or looking for food after being told no, spending a lot of time asking or talking about food, hoarding or hiding food, trying to sneak or steal food, becoming distressed when others tried to stop them asking about food or when denied a desired food that can result in verbal and physical aggression with temper tantrums, with food-related behaviour interfering with normal daily activities, such as self-care, recreation, school or work (17).

Prevention of overweight and obesity is key in the management of PWS. Excessive weight gain and its complications negatively impact on both the patient and family and incur a high degree of caregiver burden (18–23). Furthermore, there is increased mortality in PWS (24). The mean life expectancy in PWS between 1973 and 2015 was 30 years old with 70% of deaths in adults and 20% in children under 18 years old (25). Patients with PWS with unmanaged and uncontrolled obesity live on average 30 years less than those with weight control (26, 27).

As in all people with common obesity, the burden of disease in individuals with obesity due to rare genetic disorders negatively affects health-related quality of life and is associated with significant overall morbidity secondary to obesity e.g. type 2 diabetes mellitus (T2DM), hypertension, cardiovascular disease, obstructive sleep apnea, hypoventilation, pulmonary hypertension, type 2 respiratory failure (1, 5, 6, 28). Additionally, morbidity related to food seeking (e.g. accidents, choking) contribute to approximately one-third of all deaths in childhood and adolescents in PWS (25), and patients with PWS have intense impairment in both mental and physical aspects of quality of life (18, 22).

Hyperphagia is an overwhelming burden both to the individual and the family (20). Constant food seeking creates extraordinary stress on families, caregivers, and support systems. Compared with general inpatient and outpatient samples of children with complex health conditions not including PWS, family members of children with PWS showed poorer perceived quality of life, difficulties in family functioning, communication problems, increased number of conflicts, higher than average levels of depression and feelings of isolation, anger and worry (21). In the USA, caregivers of children and adults with PWS have an overall high caregiver burden, that correlated with caregiver depressed mood, anxiety, sleep and work disruption, and negative romantic relationship effect (23). Unaffected siblings of patients with PWS also describe poor quality of life, high levels of depression and anxiety, and symptoms of post-traumatic stress disorder (19, 21, 29).

### 1.3. Current Treatment of Hyperphagia and Obesity in PWS

The management of hyperphagia and obesity PWS is uniquely complex. Traditional lifestyle-based interventions, while foundational, are frequently insufficient due to the strong drive to eat and behavioural rigidity associated with the syndrome. Consequently, treatment typically involves a combination of environmental, behavioural, hormonal, and, increasingly, pharmacological strategies (1, 6, 30, 31).

Parents and caregivers must maintain a considerable level of environmental control, including supervision around food, securing food sources (e.g. locking the refrigerator and cupboards), providing reduced caloric meals and snacks, and adhering to a predictable meal schedule to reduce anxiety, and controlling access to money (1, 6, 30). While strict control of the food environment is an essential management strategy for weight regulation in PWS, they do not address the persistent hyperphagic drive. Adolescents with PWS are especially at increased risk of gaining weight for various reasons, including reduced physical activity, wish for independence resulting in unsupervised access to food, loss of routine through the changes in educational and home environments (32–34). This results in a high prevalence of obesity in PWS upon entering adulthood (82-98%) (28, 35), and the natural history is for progressive weight gain over time so that even weight maintenance is a successful outcome for interventions (36).

Where available, use of specialist PWS residential homes for adults, with strict control of the food environment, food security and encouragement of physical activity is often lifesaving to reduce severe obesity and treat obesity-related complications (37–40). However, availability of and funding for such placements is quite restricted both between and within different countries. This means that treatments that only mildly attenuate rather than normalise hyperphagia may still be extremely beneficial in PWS as it may allow residence in less specialised locations, such as continuing in the family home or in supported living or non-PWS specialist residential homes, depending on the family situation, level of intellectual disability and presence of other behavioral and psychiatric issues.

Treatment with growth hormone (GH) to improve final height, increase lipolysis and improve muscle mass has been the mainstay of treatment for children and young people with PWS for the last 2-3 decades. GH helps to improve body composition and can help maintain a healthy weight but does not affect hyperphagia (1, 6, 41, 42). Treatment with metformin is indicated for impaired glucose tolerance or T2DM, but there is little effect on weight reduction or hyperphagia (43).

Pharmacological options specifically targeting hyperphagia in PWS have been limited. Several trials of medications to reduce hyperphagia have either been unsuccessful, led to high prevalence for unacceptable side effects or have had only minimal impact (44), including endocannabinoid CB1 receptor agonist rimonabant (45), methionine aminopeptidase 2 (MetAP2) inhibitor beloranib (46), desacyl ghrelin analogue livoletide (47, 48), ghrelin O-acyl transferase (GOAT) enzyme inhibitor (49), melanocortin-4 receptor agonist setmelanotide (MC4R) (50), oxytocin and analogue carbetocin (51–55). Recently, diazoxide choline controlled release (DCCR) was approved for hyperphagia in PWS in the USA, that acts via appetite pathways in the hypothalamus. The Hyperphagia Questionnaire for Clinical Trials (HQ-CT) is a validated tool which assesses the severity of hyperphagia over the previous two weeks (**Supplementary Material**) (56), with DCCR reducing HQ-CT scores (57) and additional evidence of sustained benefit for hyperphagia and associated behavioural problems from open-label extension and placebo-controlled withdrawal studies (58–60). DCCR can however lead to side effects including peripheral oedema, hypertrichosis and hyperglycaemia, and is only currently licensed in the USA (www.vykatxr.com).

Bariatric surgery such as gastric banding, gastric bypass surgery or sleeve gastrectomy is not recommended for people with PWS because of high risk of complications and lack of evidence for long-term weight reduction with weight regain being common (27, 61).

### 1.4. GLP-1 Analogues for Non-PWS Obesity

Glucagon-like peptide-1 (GLP-1) is an incretin hormone secreted by the intestinal L-cells post-prandially. It plays a crucial role in glucose homeostasis by enhancing glucose-dependent insulin secretion (62), suppressing glucagon release and improving insulin sensitivity (63), delaying gastric emptying (64), and promoting satiety via central nervous system pathways (65). These physiological actions have been taken exploited to develop GLP-1 receptor agonists (GLP-1RAs), which were initially approved for treatment of T2DM and have since shown significant promise in obesity management (66). Semaglutide is a long-acting GLP-1RA administered by once weekly subcutaneous injection which shows resistance to degradation by dipeptidyl peptidase-4 (DPP-4) that elicits its anti-obesity effects by reducing appetite and energy intake through activation of GLP-1 receptors on the vagus nerve, hypothalamus, brainstem and brain reward pathways, in addition to its peripheral metabolic effects (67–70).

In the landmark STEP (Semaglutide Treatment Effect in People with obesity) trials sponsored by Novo Nordisk, once-weekly semaglutide at a dose of 2.4mg led to a mean weight reduction of 11.1-19.1% over 48-68 weeks (compared to placebo) in adults with obesity without T2DM (71–77), surpassing results seen with previous anti-obesity medications (see **Supplementary Table S1** for summary). Weight loss on semaglutide was notably less in adults with overweight/obesity and T2DM at 6.2% over 68 weeks (compared to placebo) (78), with lower efficacy for body weight in those with vs. without T2DM common finding across all GLP-1 analogues (79).

Semaglutide treatment reduces appetite and sweet and savoury food craving, increases satiety, improves food craving control, that are correlated with weight loss (80, 81), and improves positive mood and obesity-related quality of life (81–83). GLP-1RAs including liraglutide, dulaglutide and semaglutide may also reduce frequency of binge episodes in binge eating disorder (84–88), though there have been few placebo-controlled trials (89, 90).

Semaglutide therapy is associated with mild-moderate gastrointestinal adverse events including nausea, vomiting, abdominal discomfort, diarrhoea (up to 60-80% of patients), and increased risk of gallstones (<5%) (91, 92). Their prevalence and severity are attenuated through gradual dose titration to maintenance dose of 2.4mg once weekly over 16 weeks and/or dose reduction. Weight loss does not appear to be related to the presence of these gastrointestinal side effects (93). Discontinuation rates due to adverse events have been low in clinical trials at <10% (**Supplementary Table S1**), although this may be higher in the real world (94).

Treatment with semaglutide is generally associated with improvements in multiple cardiometabolic parameters, including glycaemic control, lipid profile, liver function tests, and blood pressure in addition to sustained weight loss, with additional potentially weight loss independent benefits on cardiovascular and renal outcomes (95–98). Currently in UK, semaglutide up to 2.4mg weekly is approved for use in obesity in adults with a body mass index (BMI) ≥30kg/m^2^ or ≥27kg/m^2^ with at least one weight-related condition such as hypertension, dyslipidaemia or T2DM.

### 1.5. GLP-1 Analogues in PWS

Early GLP-1 analogues such as exenatide and liraglutide have shown mixed results in PWS. There were some benefits in case reports in PWS (99–101), however a phase 2 clinical trial of liraglutide in children and adolescents with PWS (n=24-31) found no weight loss, though there was a suggestion of reduced hyperphagia in adolescents (102). Case reports (n=9) of adults with PWS and T2DM treated with semaglutide suggest weight loss, however hyperphagia was not assessed and doses used were generally only up to 1mg once weekly, which is lower than usual target dose for obesity of 2.4mg weekly (103–106) (see **Table 1**, that includes a summary of findings from the current study). There have been no placebo-controlled clinical trials for semaglutide in PWS. Earlier generation GLP-1 analogues and semaglutide 1.0-2.0mg once weekly appear to be well-tolerated in PWS, with gastrointestinal side effects being the most common (27, 99, 100, 102, 104, 105).

**Table 1.** Summary of case reports of semaglutide in Prader-Willi syndrome.

| Author | Year | Country | Δ weight pre-semaglutide | N | Age | Sex | BMI status | T2DM | T2DM meds. | Maximum dose | Duration | Weight baseline | BMI baseline | Δ Weight | Δ Weight | HbA1c baseline | Δ HbA1c | Adverse events |
| --- | --- | --- | --- | --- | --- | --- | --- | --- | --- | --- | --- | --- | --- | --- | --- | --- | --- | --- |
|  |  |  |  |  | (years) |  |  |  |  | (mg) | (months) | (kg) | (kg/m<sup>2</sup>) | (kg) | (%) | (mmol/mol) | (mmol/mol) |  |
| Sani | 2022 | Italy | +9.5kg ?when | 1 | 33 | M | OB | yes | metformin, insulin | 1.0 | 12 | 99.5 | 41.4 | -5.2 | -5.2 | 98 | -43 | o |
| Giménez-Palop | 2024 | Spain | ? | 4 | 25 | M | OB | yes | metformin, empagliflozin, glargine | 0.5 | 12 | 101.0 | 33.0 | 4.9 | 4.9 | 55 | -5 | o |
|  |  |  | ? |  | 40 | M | OB | yes | metformin, empagliflozin, glargine | 0.5 | 18 | 95.5 | 46.7 | -5.0 | -5.2 | 75 | -22 | o |
|  |  |  | ? |  | 43 | F | OB | yes | metformin, empagliflozin, glargine | 1.0 | 36 | 63.5 | 31.0 | -11.5 | -18.1 | 57 | -9 | o |
|  |  |  | ? |  | 31 | F | OB | yes | metformin, empagliflozin | 1.0 | 36 | 84.5 | 41.9 | -0.2 | -0.2 | 58 | 3 | o |
| Koceva | 2025 | Slovenia | +9kg ?when | 3 | 28 | F | OB | o | n/a | 1.0 | 37 | 118.0 | 55.0 | -1.0 | -0.8 | 40 | 0 | o |
|  |  |  | +34kg ?when |  | 25 | M | OB | o | n/a | 1.0 | 11 | 137.0 | 54.9 | -15.0 | -10.9 | 42 | -2 | o |
|  |  |  | ? |  | 39 | F | OB<sup>a</sup> | o<sup>b</sup> | n/a | 2.0 | 30 | 174.0 | 82.8 | -25.0 | -14.4 | 42 | ? | dyspepsia |
| Dinoi | 2025 | Italy | +13kg in prev. 27mo | 1 | 27 | M | OB | yes | o | 0.5 | 24 | 79.0 | 36.0 | -8.0 | -10.1 | 41 | -3 | o |
| Ahmed | 2026 | UK | weight loss ≤5% in prev. 7mo | 12 | 28.3 ± 10.1 (18-51) | 10F, 2M | OW (1), OB (11) | yes (9), o (3) | metformin (8), SGLT2i (1), insulin (3), DPP4i (1), pioglitazone (2), liraglutide (1), oral semaglutide (1) | 1.0 (3), 1.7 (3), 2.4 (6)<sup>c</sup> | 20.3 ± 9.4 (8.7-36.1) | 107.0 ± 22.1 (55.2-135.8) | 46.2 ± 8.2 (28.2-55.8) | -3.8 ± 11.8 (-34.5 to +8.6) | -3.1 ± 9.9 (-27.0 to +7.4) | 62.9 ± 26.0 (39-106) | -6.9 ± 33.4 (-63 to +64) | diarrhoea (2), abdo. pain (2), dizziness (1), low mood (1), sleep disturb. (1), emotional lability (1) |
|  |  |  | weight loss >5% in prev. 7mo | 2 | 26.0 ± 8.5 (20-32) | 1F, 1M | OB (2) | yes (2) | metformin (2), insulin (1) | 0.5 (1), 1.0 (1) | 19.5 ± 19.5 (5.7-33.3) | 101.5 ± 35.1 (76.6-126.3) | 37.8 ± 8.4 (31.9-43.7) | 3.2 ± 14.8 (-7.3 to +13.7) | 0.7 ± 14.4 (-9.5 to +10.8) | 39.5 ± 0.7 (39-40) | -1.5 ± 0.7 (-2 to -1) | diarrhoea (1) |

<sup>a</sup> Underwent gastric banding and subsequently Roux-en-Y gastric bypass surgery 10 years prior to starting semaglutide with initial weight loss and subsequent weight regain. <sup>b</sup> No T2DM at baseline, but developed T2DM on treatment with semaglutide. <sup>c</sup> Three participants still undergoing up-titration of semaglutide. For summary data from the current paper Ahmed (2026), (n) indicates number of individuals, values given as mean ± SD (range). To convert HbA1c (mmol/mol) to (%) use the formula: % = (0.09148 × mmol/mol) + 2.15. Abbreviations: ?, unknown; abdo., abdominal; BMI, body mass index; disturb., disturbance; DPP4i, Dipeptidyl peptidase- 4 inhibitor; F, female; HbA1c, glycated haemoglobin; M, male; meds., medications; mo, months; n/a, not applicable; o, no; OB, obesity; OW, overweight; prev., previous; SD, standard deviation; SGLT2i, sodium-glucose co-transporter 2 inhibitor; T2DM, type 2 diabetes mellitus.

Early reports suggest that semaglutide and newer incretin agents such as the dual GLP-1/GIP analogue tirzepatide may be effective for weight loss in other genetic obesity disorders involving hypothalamic dysfunction, including variants in the melanocortin-4 receptor (*MC4R*) gene (107, 108) and Alstrom syndrome (109). Furthermore, similar trajectories of weight loss have been seen with semaglutide in adolescents with obesity and dominant or known risk gene variants for obesity, compared to those with carriers of variants in recessive genes and those without identified genetic causes, including *RAI1*, *ALMS1, IFT172, LEPR, MC4R, MKKS* and *PCSK1* genes that are involved in hypothalamic regulation of appetite (110).

Additional investigation of the efficacy and safety of GLP-1 analogues are therefore needed in PWS both through observational and placebo-controlled studies (111).

### 1.6. Hypotheses and Aims

The primary aim of this retrospective observational cohort study was to evaluate the effects of semaglutide in adults with PWS attending the genetic obesity clinic at Hammersmith Hospital who received semaglutide as part of routine clinical care. The analysis aimed to assess changes in weight and monthly weight trajectory, expressed as both absolute and percentage change, with comparison to longitudinal weight trends prior to initiation of semaglutide. Additional outcomes of interest include changes in hyperphagia, measured using the Hyperphagia Questionnaire for Clinical Trials (HQ-CT), glycaemic control as assessed by glycated haemoglobin (HbA1c), lipid parameters including low-density lipoprotein cholesterol (LDL-cholesterol), high-density lipoprotein cholesterol (HDL-cholesterol), total/HDL cholesterol ratio (total /HDL-cholesterol), and liver biochemistry markers, specifically alanine transaminase (ALT) and aspartate aminotransferase (AST). A secondary aim was to characterise the safety and tolerability of semaglutide in adults with PWS by assessing the frequency and nature of adverse events, as well as the need for dose adjustments or treatment discontinuation during follow-up.

It was hypothesised that treatment with semaglutide will lead to clinically meaningful weight reduction, although probably with a smaller magnitude of effect compared with that observed in prior studies of individuals with non-genetic obesity, or at least weight maintenance. In addition, semaglutide was hypothesised to improve hyperphagia and food-seeking behaviours, glycaemic control, lipid profile, and markers of metabolic associated steatotic liver disease. It was further hypothesised that semaglutide would be well tolerated in this population, with a safety profile comparable to that reported in studies of non-genetic obesity.

## 2. MATERIALS AND METHODS

### 2.1. Study Design

This was a retrospective, observational cohort study conducted at a UK academic hospital centre specialist adult PWS clinic that reviewed clinical data from adult patients with PWS who were treated with semaglutide as part of routine clinical care.

### 2.2. Participants and Inclusion/Exclusion Criteria

Eligible participants were male or female adults aged ≥18 years with genetically confirmed PWS (by *SNRPN* gene methylation analysis), with overweight/obesity, defined as BMI ≥25 kg/m^2^, and with or without T2DM, commencing treatment with subcutaneous semaglutide in the clinic. Additional inclusion criteria included: documented clinical follow-up for at least one time point after treatment initiation, and availability of weight data and relevant clinical outcomes. Participants who had lost more than 5% in body weight in the seven months prior to starting semaglutide were excluded from analyses of efficacy, but were included in assessment of side effects. Permission to access this clinical data was obtained as part of a PWS clinic service evaluation approved by Imperial College Healthcare National Health Service Trust.

Patients received semaglutide 2.4mg (or maximum-tolerated dose), subject to clinical guidelines and drug availability. Semaglutide was administered once weekly as subcutaneous injections in the abdomen, thigh or upper arm. Semaglutide was initiated at 0.25mg and up titrated at a minimum of every four weeks to 0.5mg, then 1mg, then 1.7mg and finally 2.4mg or until maximum tolerated dose. During dose escalation most clinic visits were held by telephone or video rather than face-to-face. Participants started on semaglutide between June 2022 and July 2025, and data collection was until December 2025. All eligible patients during the study period were included.

### 2.3. Data Collection

Data were extracted from electronic health records at all hospital clinic attendances: at up to three time points before treatment initiation including baseline, and all timepoints after starting treatment. For each time point, the following variables were recorded were available: weight (kg) to calculate BMI (kg/m^2^); hyperphagia (HQ-CT) and food environment (FSZ) questionnaires; metabolic parameters (measured in Dept. of Clinical Biochemistry, Imperial College Healthcare NHS Trust, London, UK): HbA1c, non-fasting lipid profile (total-cholesterol, LDL-cholesterol, HDL-cholesterol, total/HDL-cholesterol ratio), liver enzymes ALT and AST); safety data (reported side effects, dose modifications, discontinuation due to tolerability). ALT, AST, platelet count and age were used to calculate FIB-4 score indicating risk of hepatic fibrosis using formula [age (years) x AST (IU/L)] / [platelet count (x10^9^/L) / √ ALT (IU/L)], with <1.45 having negative predictive value of 90% for advanced fibrosis and score >3.25 having 97% specificity and positive predictive value of 65% for advanced fibrosis (112).

### 2.4. Hyperphagia Questionnaires

The HQ-CT was completed by carers/parents to assesses hyperphagia over the previous two weeks, comprising nine questions each scored from 0-4, with total scores ranging from 0-36, with a higher score indicating greater severity of hyperphagia (**Supplementary Material**) (56). HQ-CT sub-scales assessed hyperphagia behaviour (0-16), drive (0-12) and severity (0-8), respectively measuring the frequency and intensity of food-seeking actions, emotional reactions to food restriction, and preoccupation with food.

Hyperphagia over the previous year was also measured prior to initiation of semaglutide using the Dykens Hyperphagia Questionnaire, comprising 13 questions each scored from 1-5, with total scores ranging from 11-55, with a higher score indicating greater severity of hyperphagia (17).

### 2.5. Food Safe Zone Questionnaire

The Food Safe Zone questionnaire (FSZ) was completed by carers/parents as a measure of food security and how strictly the food environment is controlled, comprising 20 questions, with scores ranging from 0-80, with a higher score indicating a more strictly controlled food environment (113).

### 2.6. Statistical Analysis

All data were statistically processed using GraphPad Prism-10 (GraphPad Software Inc., Boston, MA, USA), and normality assessed using Kolmogorov-Smirnov test. P values <0.05 were taken as statistically significant. Where data was available across three time points, namely visit previous to baseline (pre-treatment), baseline (treatment initiation), and post-visit, statistical analysis used repeated measures ANOVA with mixed model analysis (to account for missing data) using post-hoc Fisher LSD test, and paired t-test for comparison of change in outcome variable pre-visit vs. baseline compared to baseline vs. post-visit. Where graphs show data across two time points, namely baseline (treatment initiation) and post-visit, paired t-test was performed. Standardised effect sizes were also reported as Cohen’s d to avoid over-reliance on interpretation of P-values with the small sample size.

## 3. RESULTS

### 3.1. Demographic Data

**Table 2** summarises the baseline demographic and clinical characteristics of participants prior to semaglutide initiation. Twelve participants who had experienced ≤5% weight loss in the preceding seven months were included in the primary analysis: mean ± SD age 28.3 ± 10.1 years (range 18-51), 10 (83.3%) female, all with overweight/obesity, nine (75.0%) with T2DM, with four of these having poor glycaemic control with HbA1c >58 mmol/mol or >7.5%. For those with T2DM, the medications used were as follows: four metformin only (one of whom started metformin at the same time as semaglutide); one metformin and DPP4 inhibitor; one metformin, thiazolidinedione and insulin (22 units daily); one metformin, sodium-glucose co-transporter 2 inhibitor and liraglutide 1.8mg subcutaneous once daily; one metformin, thiazolidinedione, and oral semaglutide 3mg once daily (equivalent to 0.11mg subcutaneous semaglutide); one insulin only (50 units daily).

**Table 2.**
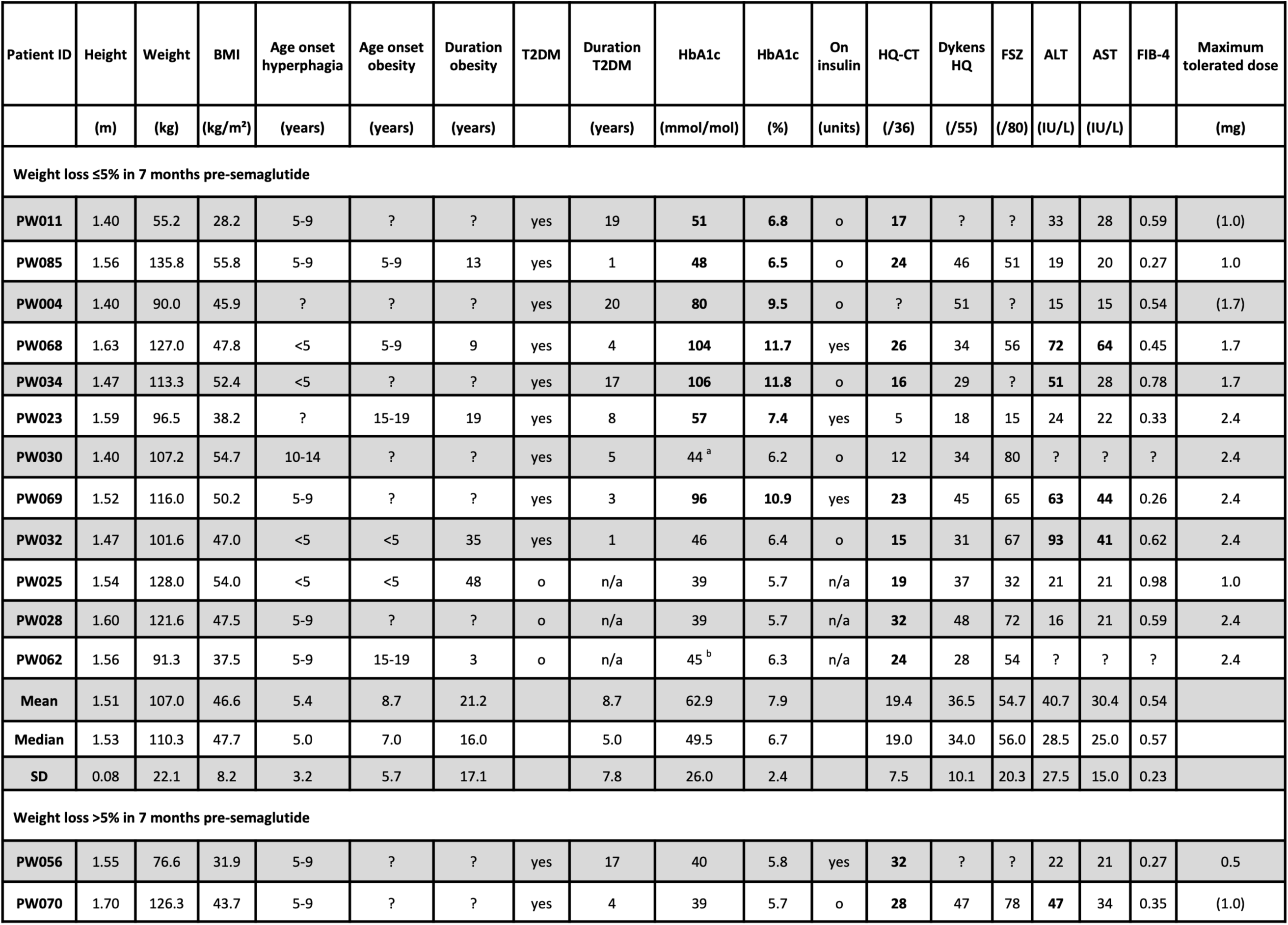
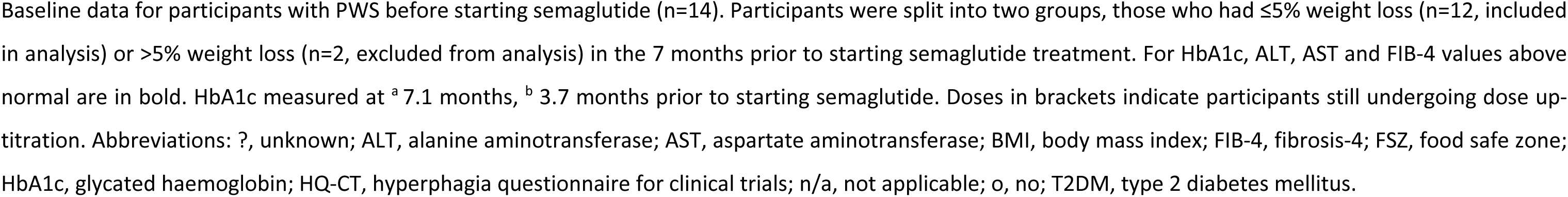
Baseline demographic and clinical characteristics of participants.

| Patient ID | Height | Weight | BMI | Age onset hyperphagia | Age onset obesity | Duration obesity | T2DM | Duration T2DM | HbA1c | HbA1c | On insulin | HQ-CT | Dyken's HQ | FSZ | ALT | AST | FIB-4 | Maximum tolerated dose |
| --- | --- | --- | --- | --- | --- | --- | --- | --- | --- | --- | --- | --- | --- | --- | --- | --- | --- | --- |
|  | (m) | (kg) | (kg/m<sup>2</sup>) | (years) | (years) | (years) |  | (years) | (mmol/mol) | (%) | (units) | (/36) | (/55) | (/80) | (IU/L) | (IU/L) |  | (mg) |
| Weight loss ≤5% in 7 months pre-semaglutide |  |  |  |  |  |  |  |  |  |  |  |  |  |  |  |  |  |  |
| PW011 | 1.40 | 55.2 | 28.2 | 5-9 | ? | ? | yes | 19 | 51 | 6.8 | o | 17 | ? | ? | 33 | 28 | 0.59 | (1.0) |
| PW085 | 1.56 | 135.8 | 55.8 | 5-9 | 5-9 | 13 | yes | 1 | 48 | 6.5 | o | 24 | 46 | 51 | 19 | 20 | 0.27 | 1.0 |
| PW004 | 1.40 | 90.0 | 45.9 | ? | ? | ? | yes | 20 | 80 | 9.5 | o | ? | 51 | ? | 15 | 15 | 0.54 | (1.7) |
| PW068 | 1.63 | 127.0 | 47.8 | <5 | 5-9 | 9 | yes | 4 | 104 | 11.7 | yes | 26 | 34 | 56 | 72 | 64 | 0.45 | 1.7 |
| PW034 | 1.47 | 113.3 | 52.4 | <5 | ? | ? | yes | 17 | 106 | 11.8 | o | 16 | 29 | ? | 51 | 28 | 0.78 | 1.7 |
| PW023 | 1.59 | 96.5 | 38.2 | ? | 15-19 | 19 | yes | 8 | 57 | 7.4 | yes | 5 | 18 | 15 | 24 | 22 | 0.33 | 2.4 |
| PW030 | 1.40 | 107.2 | 54.7 | 10-14 | ? | ? | yes | 5 | 44<sup>a</sup> | 6.2 | o | 12 | 34 | 80 | ? | ? | ? | 2.4 |
| PW069 | 1.52 | 116.0 | 50.2 | 5-9 | ? | ? | yes | 3 | 96 | 10.9 | yes | 23 | 45 | 65 | 63 | 44 | 0.26 | 2.4 |
| PW032 | 1.47 | 101.6 | 47.0 | <5 | <5 | 35 | yes | 1 | 46 | 6.4 | o | 15 | 31 | 67 | 93 | 41 | 0.62 | 2.4 |
| PW025 | 1.54 | 128.0 | 54.0 | <5 | <5 | 48 | o | n/a | 39 | 5.7 | n/a | 19 | 37 | 32 | 21 | 21 | 0.98 | 1.0 |
| PW028 | 1.60 | 121.6 | 47.5 | 5-9 | ? | ? | o | n/a | 39 | 5.7 | n/a | 32 | 48 | 72 | 16 | 21 | 0.59 | 2.4 |
| PW062 | 1.56 | 91.3 | 37.5 | 5-9 | 15-19 | 3 | o | n/a | 45<sup>b</sup> | 6.3 | n/a | 24 | 28 | 54 | ? | ? | ? | 2.4 |
| Mean | 1.51 | 107.0 | 46.6 | 5.4 | 8.7 | 21.2 |  | 8.7 | 62.9 | 7.9 |  | 19.4 | 36.5 | 54.7 | 40.7 | 30.4 | 0.54 |  |
| Median | 1.53 | 110.3 | 47.7 | 5.0 | 7.0 | 16.0 |  | 5.0 | 49.5 | 6.7 |  | 19.0 | 34.0 | 56.0 | 28.5 | 25.0 | 0.57 |  |
| SD | 0.08 | 22.1 | 8.2 | 3.2 | 5.7 | 17.1 |  | 7.8 | 26.0 | 2.4 |  | 7.5 | 10.1 | 20.3 | 27.5 | 15.0 | 0.23 |  |
| Weight loss >5% in 7 months pre-semaglutide |  |  |  |  |  |  |  |  |  |  |  |  |  |  |  |  |  |  |
| PW056 | 1.55 | 76.6 | 31.9 | 5-9 | ? | ? | yes | 17 | 40 | 5.8 | yes | 32 | ? | ? | 22 | 21 | 0.27 | 0.5 |
| PW070 | 1.70 | 126.3 | 43.7 | 5-9 | ? | ? | yes | 4 | 39 | 5.7 | o | 28 | 47 | 78 | 47 | 34 | 0.35 | (1.0) |

Baseline data for participants with PWS before starting semaglutide (n=14). Participants were split into two groups, those who had $\leq 5\%$ weight loss (n=12, included in analysis) or $> 5\%$ weight loss (n=2, excluded from analysis) in the 7 months prior to starting semaglutide treatment. For HbA1c, ALT, AST and FIB-4 values above normal are in bold. HbA1c measured at <sup>a</sup> 7.1 months, <sup>b</sup> 3.7 months prior to starting semaglutide. Doses in brackets indicate participants still undergoing dose up- titration. Abbreviations: ?, unknown; ALT, alanine aminotransferase; AST, aspartate aminotransferase; BMI, body mass index; FIB-4, fibrosis-4; FSZ, food safe zone; HbA1c, glycated haemoglobin; HQ-CT, hyperphagia questionnaire for clinical trials; n/a, not applicable; o, no; T2DM, type 2 diabetes mellitus.

All but one participant had at least one obesity-related complications including hypertension (n=3, 25.0%), hyperlipidaemia (n=5, 41.7%), obstructive sleep apnoea (n=7, 58.3%), metabolic dysfunction-associated steatotic liver disease (n=7, 58.3%), obesity-related mobility problems (n=3, 25.0%), current pre-diabetes (n=1, 8.3%), microalbuminuria (n=5, 41.7%). All had genetically confirmed PWS, with an equal genotype ratio of paternal chromosome 15q11-13 deletion (n=5) vs. maternal uniparental disomy (mUPD, n=3) or mUPD/imprinting defect (n=2) where known, with two having unknown PWS genotype. All participants with available data (n=11) had a HQ-CT questionnaire score ≥5/36, and 81.8% had HQ-CT ≥13/36 indicating at least moderate hyperphagia. Ten (83.3%) patients lived in the family home, one of whom moved to supported living 8 months after starting semaglutide, with the other two (16.7%) living in PWS specialist residential homes. Seven (58.3%) had received growth hormone treatment during childhood, with four (33.3%) on growth hormone and eight (75.0%) on sex steroid replacement at baseline. Only two participants (16.7%) were taking psychotropic medications, both of which have been associated with weight gain (risperidone, olanzapine), with one increasing the dose 6 months into treatment with semaglutide.

Two participants who had lost >5% of body weight during this period were excluded and are presented separately. Their ages were in range 20-34 years, one was female, one had paternal chromosome 15q11-13 deletion and one unknown PWS genotype. Only one was living in the family home, both had T2DM receiving metformin alone or metformin and insulin (14 units daily), both had obstructive sleep apnoea, and one had hypertension, hyperlipidaemia or microalbuminuria.

### 3.2. Semaglutide Dose and Side Effects

In the 12 patients who had not lost weight prior to starting semaglutide, 78.6% (12/14) had reached their maximum tolerated dose of semaglutide by the end of follow-up (two still being up-titrated), with 60.0% of these (6/10) tolerating the maximum dose (2.4mg once weekly), with median semaglutide dose 2.4mg once weekly (1.0-2.4), for a median follow-up of 17.2 months (range 8.7-36.1), with median time on highest dose 6.5 months (range 2.6-29.4) (**Table 4**).

In the whole cohort (including the two participants with weight loss prior to starting semaglutide), 78.6% (11/14) had reached their maximum tolerated dose of semaglutide by the end of follow-up (three still being up-titrated), with 54.5% of these (6/11) tolerating the maximum dose (2.4mg once weekly), for a median follow-up time of 16.6 months (range 5.7-36.1), with median time on highest dose 6.5 months (range 1.8-29.4) (**Table 4**).

Less than half (6/14, 42.9%) of participants experienced at least one side effect, with only two participants having more than one side effect (**Table 4**, **Figure 4**). The most common side effect was diarrhoea (3/14, 21.4%, one of whom had newly diagnosed ulcerative colitis), followed by abdominal pain (2/14, 14.3%), and then dizziness, sleep disturbance, low mood and emotional lability (1/14, 7.1% each). No cases of nausea, vomiting or constipation were reported. Overall, 4/14 (28.6%) had at least one gastrointestinal side effect.

Among the six participants who experienced side effects on semaglutide, 16.6% (1/6) required a temporary delay in dose escalation, and 66.6% (4/6) required a dose reduction. Importantly, no participants discontinued semaglutide treatment due to side effects.

### 3.3. Weight Change

In those 12 participants who had not been losing weight prior to semaglutide initiation, median follow-up duration on semaglutide was 17.2 months, median duration on maximum semaglutide dose was 6.5 months **(Table 3)**. Two participants were still uptitrating their semaglutide dose, with six (50.0%) of participants reaching and tolerating the maximum semaglutide dose 2.4mg once weekly **(Table 3)**.

**Table 3.**
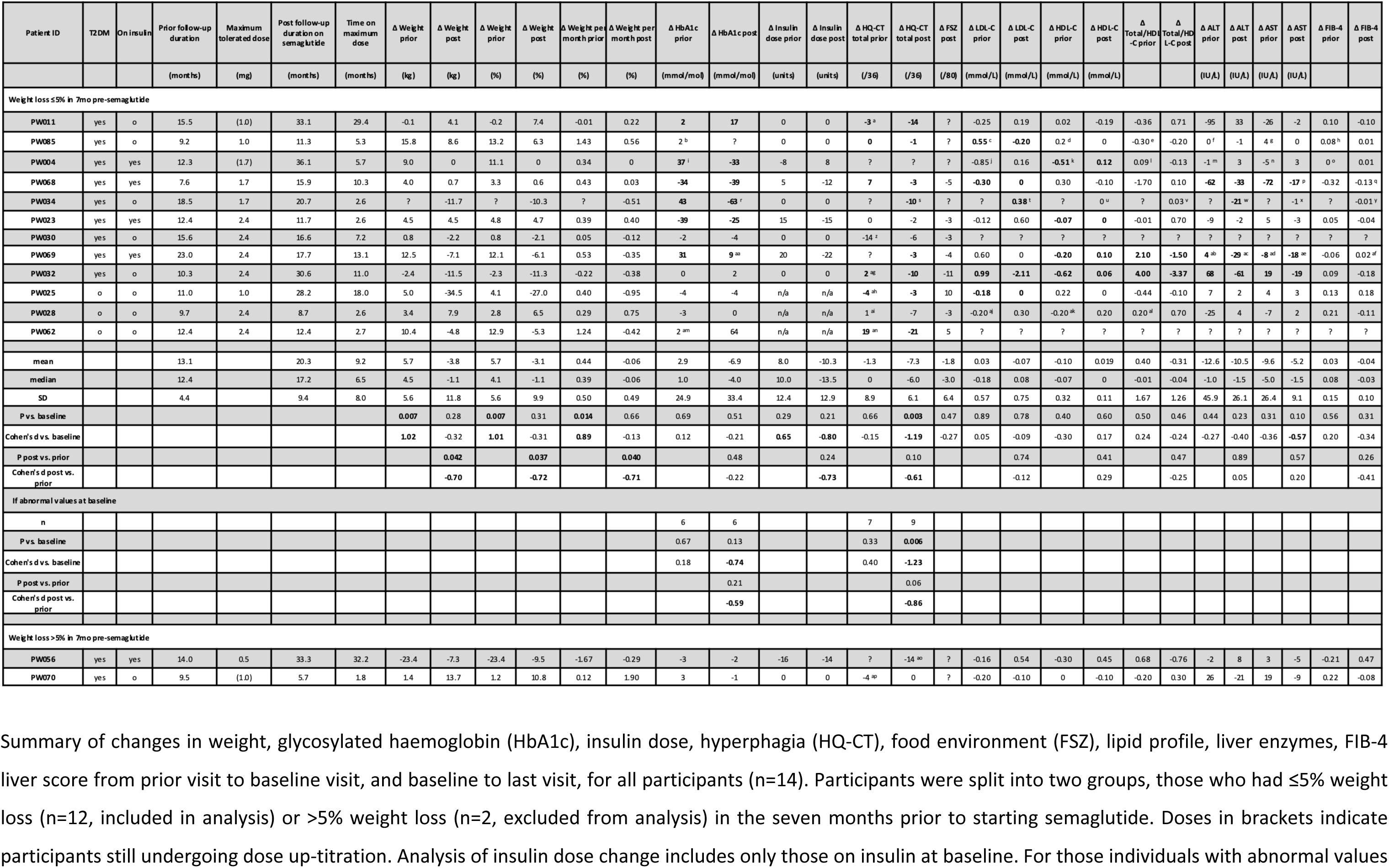
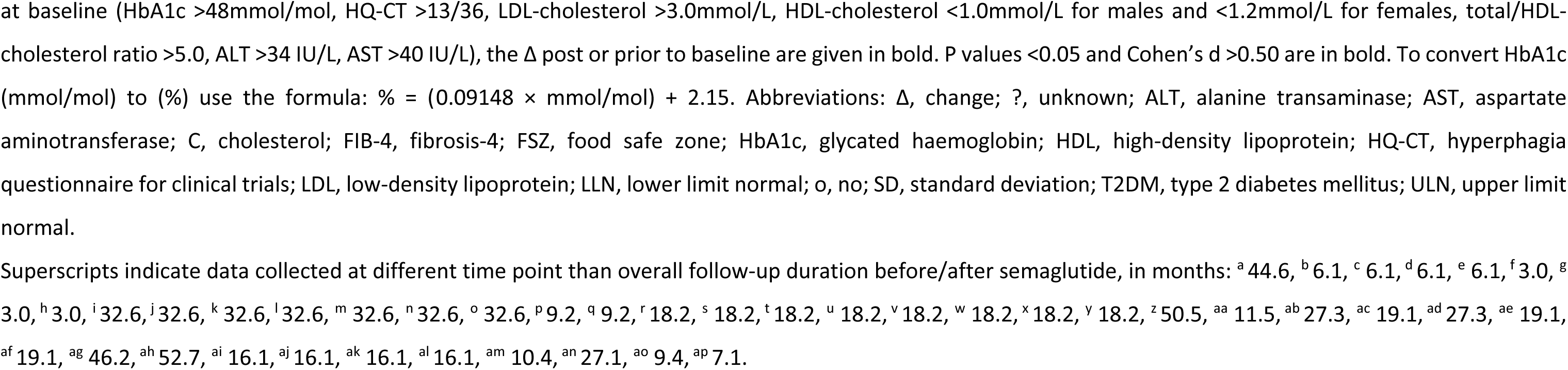
Summary of changes in outcomes prior to and after starting semaglutide.

*Prior* to semaglutide initiation, most participants were gaining weight, with mean ± SD weight change +5.7 ± 5.6% from median of 12.4 months before semaglutide initiation to baseline (n=11, P=0.007, Cohen’s d +1.01, excluding PW034 who did not have a recorded weight at the prior visit) (**Table 3**, **Figure 1A**, **Figure 1C**). Following initiation of semaglutide over median follow-up of 17.2 months, there was no statistically significant change in weight, with mean ± SD weight change -3.1 ± 9.9% (n=12, P=0.31, d -0.31). However, comparison of weight trajectories *before vs. after* treatment demonstrated a significant attenuation of pre-treatment weight gain after semaglutide initiation with effect size -8.1 ± 11.2% body weight (n=11, P=0.037, d -0.72) (**Table 3**, **Figure 1C**). Similar results were seen, when comparing pre- vs. post-semaglutide weight change, as absolute weight change in kg (n=11, P=0.042, d -0.70) (**Table 3**), or as percentage weight change per month (n=11, P=0.040, d -0.71) (**Table 3**, **Figure 1D**).

**Figure 1.**
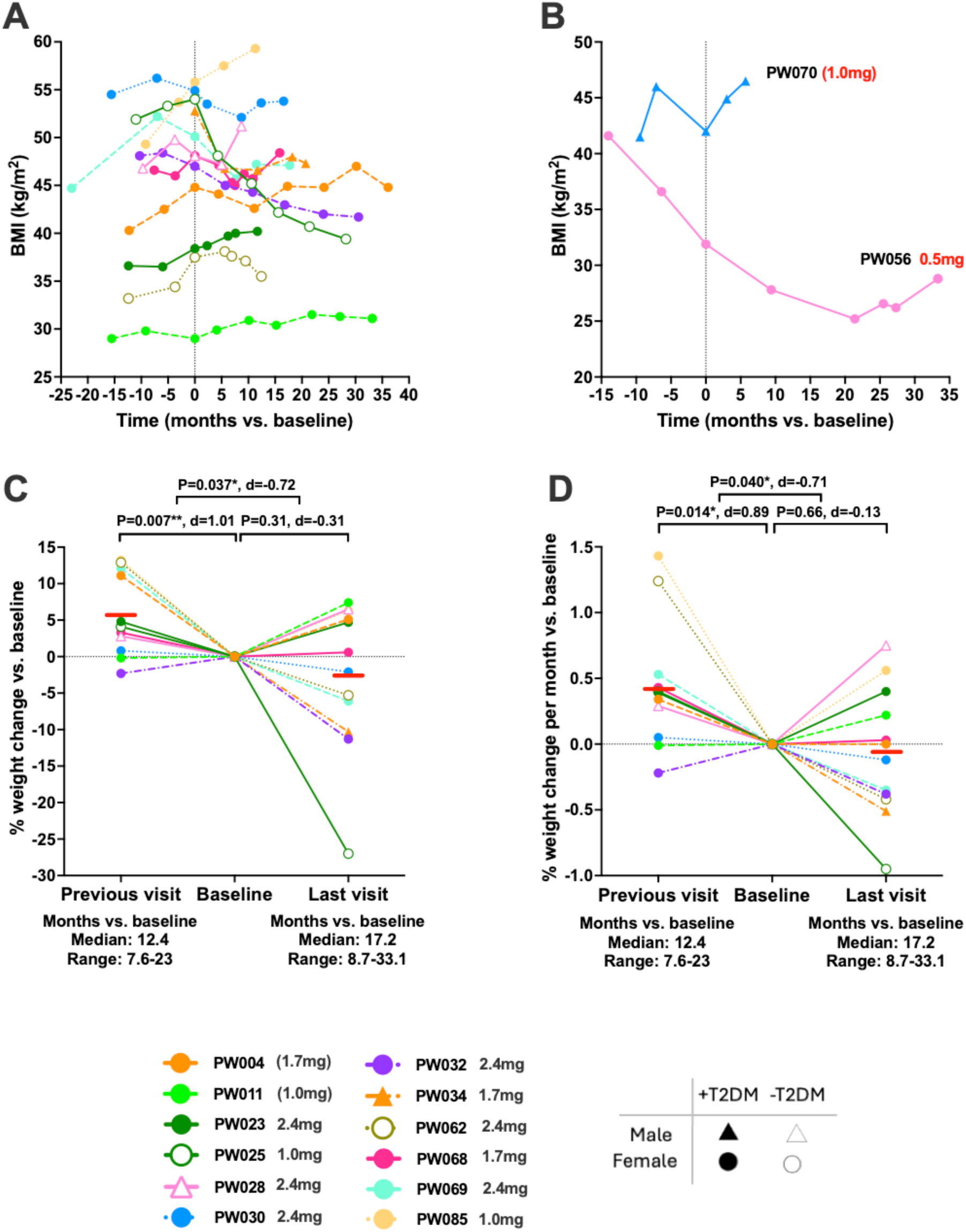
Changes in BMI and weight before and after starting semaglutide in adults with PWS. Each graph displays individual participant trajectories, both before and following treatment with semaglutide. Doses in brackets indicate participants still undergoing dose up-titration. Graphs show (A) change in BMI over time for those with ≤ 5% weight loss over seven months before starting semaglutide (n=12), with latest drug dose noted for each participant at bottom of figure; (B) change in BMI over time for those with > 5% weight loss over seven months before starting semaglutide (n=2), with latest drug dose noted for each participant to right of graph lines in red; (C) percentage weight change vs. baseline, and (D) percentage weight change per month vs. baseline, at visits prior to starting drug and at last follow-up (participants from A, n=12). Mean values are indicated by red lines. Median and range of time (in months) from baseline are given below x-axis for visits. Statistical comparisons in (C,D) used repeated measured ANOVA with mixed model analysis using post-hoc Fisher LSD test and paired t-test for comparison of change in outcome variable pre- visit vs. baseline vs. baseline vs. post-visit, with P-values and Cohen’s d effect sizes * P<0.05, ** P<0.01. Abbreviations: ANOVA, repeated-measures analysis of variance; BMI, body mass index; T2DM, type 2 diabetes mellitus.

After receiving semaglutide, the participants without T2DM experienced a numerically greater reduction (P=0.47, d -0.51) in body weight (n=3, mean ± SD -8.6 ± 17.0%) compared to those with T2DM (n=9, -1.2 ± 6.9%), despite similar % weight gain prior to starting semaglutide (+6.6 ± 5.5 vs. +5.4 ± 6.0), a shorter duration of follow-up (median 12.4 vs. 17.7 months), and similar median semaglutide dose of 2.4mg once weekly. Indeed, the largest individual reduction in body weight (-27.0% over 28.2 months) occurred in a participant without T2DM (PW025) and despite tolerating only semaglutide 1.0 weekly **(Table 3)**.

For the weight trajectories of the two participants excluded from analysis as they had lost >5% of body weight in the seven months prior to treatment initiation, one appeared to have some slowing of weight loss (PW056, tolerating up to 0.5mg weekly), while the other regained weight (PW070, still on 1.0mg once weekly during up-titration) (**Figure 1B**).

### 3.4. Hyperphagia Questionnaire

The change in total HQ-CT score (out of 36) *prior* to treatment was not significant (n=9, mean ± SD -1.3 ± 8.9, P=0.66, d -0.15), whereas there was a significant reduction in total HQ-CT score *following* semaglutide treatment (n=11, -7.3 ± 6.1, P=0.003, d -1.19). Although the difference in change of total HQ-CT between the pre- and post-treatment periods did not reach statistical significance, there was a moderate effect size (n=9, -8.3 ± 13.6, P=0.10, d -0.61) (**Table 3**, **Figure 2A)**.

**Figure 2.**
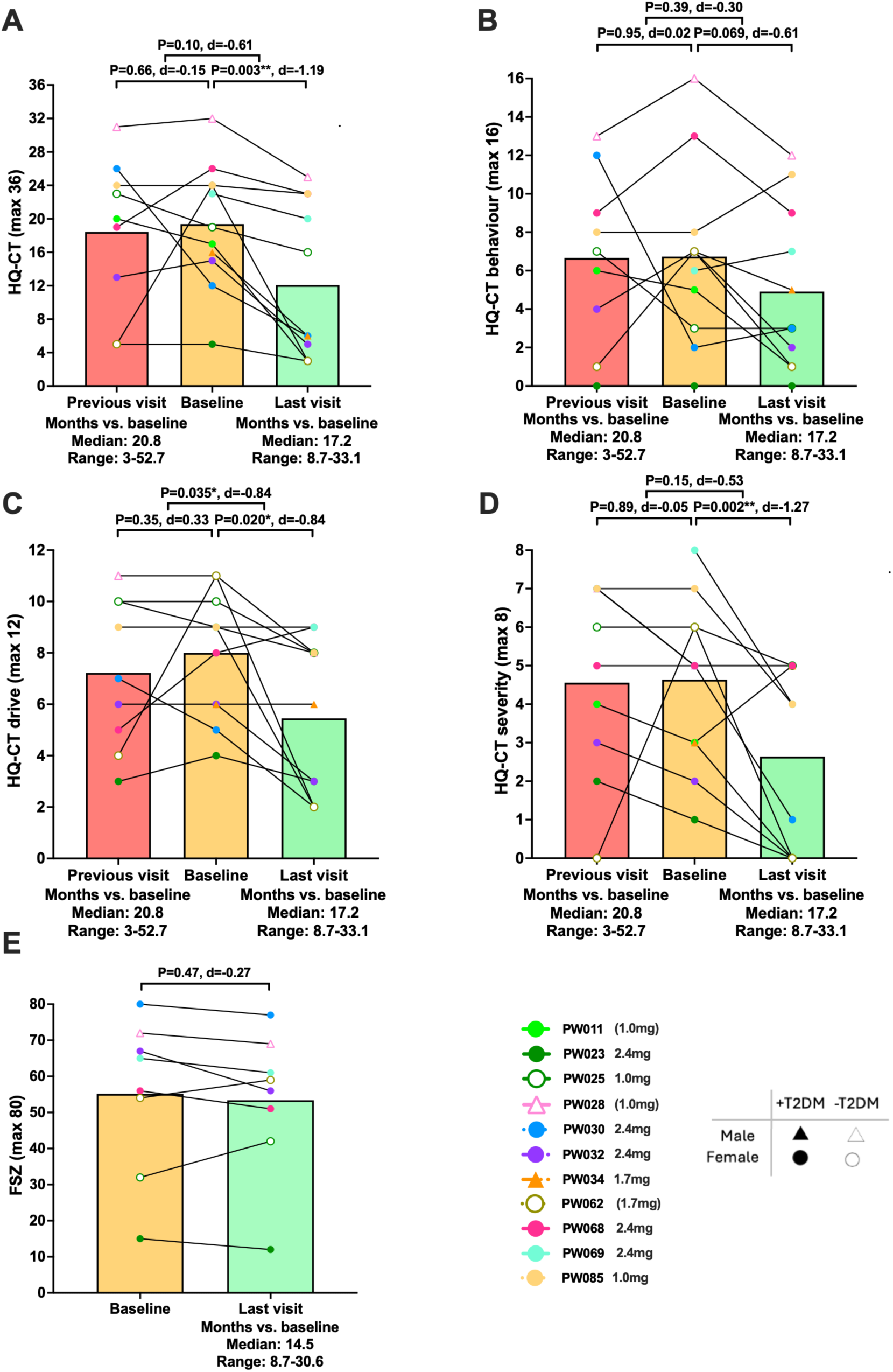
Hyperphagia questionnaire score outcomes in adults with PWS treated with semaglutide. All graphs depict individual level data across up to three time points (visit previous to baseline (pre-treatment), baseline (treatment initiation), and last available follow-up), for (A) HQ-CT total scores, n=11; (B) HQ-CT behaviour scores, n=11; (C) HQ-CT drive scores, n=11; (D) HQ-CT severity scores, n=11; (E) FSZ scores, n=8, in those with ≤ 5% weight loss over seven months before starting semaglutide. Each graph displays individual participant trajectories, with bar representing mean at each time point, with semaglutide doses noted for each participant, and doses in brackets indicating participants still undergoing dose up-titration. Median and range of time (in months) from baseline are given below x-axis for visits. Statistical analysis in (A-D) used repeated measures ANOVA with mixed model analysis using post-hoc Fisher LSD test, and paired t-test for comparison of change in outcome variable pre-visit vs. baseline and baseline vs. post-visit, and in (E) paired t-test, with P-values and Cohen’s d effect sizes displayed: * P<0.05, ** P<0.01. Abbreviations: ANOVA, repeated-measures analysis of variance; FSZ, food safe zone; HQ-CT, hyperphagia questionnaire for clinical trials; T2DM, type 2 diabetes mellitus.

Analysis of HQ-CT sub-scales (behaviour, drive, severity) demonstrated similar patterns. HQ-CT behaviour scores (out of 16) did not change significantly *prior* to treatment (n=9, +0.1 ± 4.8, P=0.95, d +0.02) and showed a trend for reduction *following* semaglutide treatment (n=11, -1.8 ± 3.0, P=0.069, d -0.61), with no significant difference between pre- and post-treatment periods (n=9, -2.2 ± 7.3, P=0.39, d -0.30) (**Table 3**, **Figure 2B**). HQ-CT drive scores (out of 12) showed no significant change *prior* to treatment (n=9, +0.9 ± 2.7, P=0.35, d +0.33), but decreased significantly *following* semaglutide treatment (n=11, -2.5 ± 3.0, P=0.020, d -0.84), with a significant difference between the pre- and post-treatment periods (n=9, -4.0 ± 4.7, P=0.035, d -0.84) (**Table 3**, **Figure 2C**). Similarly, severity scores (out of 8) were stable *prior* to treatment (n=9, -0.1 ± 2.4, P=0.89, d - 0.05), but decreased significantly *following* semaglutide treatment (n=11, -2.4 ± 1.9, P=0.002, d -1.27), although the difference between the pre- and post-treatment periods was not statistically significant, there was a moderate effect size (n=9, -2.1 ± 4.0, P=0.15, d -0.53) (**Table 3**, **Figure 2D**).

In the nine participants with total HQ-CT score over 13/36 at baseline, the change in total HQ-CT score *prior* to treatment was not significant (n=7, mean ± SD +3.1 ± 7.9, P=0.33, d +0.40), whereas there was a significant reduction in total HQ-CT score *following* semaglutide treatment (n=9, -8.0 ± 6.5, P=0.006, d -1.23). Although the difference in change in total HQ-CT between the pre- and post-treatment periods did not reach statistical significance, there was a large effect size (n=7, -11.6 ± 13.5, P=0.06, d -0.86) (**Table 3**, **Figure 2A**).

In the five participants with total HQ-CT score over 22/36 at baseline, the change in total HQ-CT score *prior* to treatment was not significant but with a moderate increasing effect size (n=4, mean ± SD +6.8 ± 8.7, P=0.22, d +0.77), and although not reaching statistical significance, there was a large effect size for decrease in HQ-CT *following* semaglutide treatment (n=5, -7.0 ± 8.1, P=0.13, d -0.86). Similarly, the difference in change in total HQ-CT between the pre- and post-treatment periods did not reach statistical significance, but there was a large effect size (n=4, -14.8 ± 17.3, P=0.19, d -0.85) (**Table 3**, **Figure 2A**).

For the two participants excluded from overall analysis because of pre-initiation weight loss, PW056 showed a reduction in HQ-CT total score from 32/36 to 18/36 at 9.4 months *following* semaglutide treatment; whilst for PW070, HQ-CT total score had increased from 28/36 to 32/36 over the 7.1 months *before* starting semaglutide, and was stable after 5.7 months on semaglutide at 28/36 (**Table 3**).

### 3.5. Food Safe Zone and Potential Confounds

FSZ scores did not change significantly *following* semaglutide initiation (n=8, -1.8 ± 6.4, P=0.47, d -0.27) (**Table 3**, **Figure 2E**), indicating that improvements in hyperphagia and weight were unlikely to be explained by changes in their food environment, although scores were missing for four participants.

Only 4/14 of the patients had confounders over the course of their treatment with semaglutide that may have affected weight and glycaemic control, including moving residence from family home to supported living (1/14, 7.1%, PW032) and switching schools (1/14, 7.1%, PW069), small dose increase in risperidone (1/14, 7.1%, PW030), and commencement of metformin (2/14, 14.3%, PW034 at baseline; PW069 at 17 months into 17.4 months treatment with semaglutide, but HbA1c was analysed prior to starting metformin) (**Table 4**).

**Table 4.** Summary of adverse events and tolerability.

| Patient ID | T2DM | Maximum tolerated dose (mg) | Total duration semaglutide treatment (months) | Duration on maximum tolerated dose (months) | Adverse events | Dose at adverse event (mg) | Dosing outcome with adverse event |
| --- | --- | --- | --- | --- | --- | --- | --- |
| **Weight loss ≤5% in 7 months pre-semaglutide** |  |  |  |  |  |  |  |
| PW011 | yes | (1.0) | 33.6 | 29.4 | o | n/a | n/a |
| PW085 | yes | 1.0 | 9.6 | 5.3 | yes | 0.5 | delay increase to 1.0 |
| PW004 | yes | (1.7) | 35.9 | 5.7 | o | n/a | n/a |
| PW068 | yes | 1.7 | 14.4 | 10.3 | yes | 2.4 | decrease to 1.7 |
| PW034 | yes | 1.7 | 20.5 | 2.6 | yes | 2.4 | decrease to 1.7 |
| PW023 | yes | 2.4 | 11.9 | 2.6 | yes | 1.7 | none |
| PW030 | yes | 2.4 | 16.3 | 7.2 | o | n/a | n/a |
| PW069 | yes | 2.4 | 17.4 | 13.1 | o | n/a | n/a |
| PW032 | yes | 2.4 | 29.6 | 11.0 | o | n/a | n/a |
| PW025 | o | 1.0 | 27.5 | 18.0 | yes | 1.7 | decrease to 1.0 |
| PW028 | o | 2.4 | 8.8 | 2.6 | o | n/a | n/a |
| PW062 | o | 2.4 | 10.6 | 2.7 | o | n/a | n/a |
| **Weight loss >5% in 7 months pre-semaglutide** |  |  |  |  |  |  |  |
| PW056 | yes | 0.5 | 33.2 | 32.2 | yes | 1.0 | decrease to 0.5 |
| PW070 | yes | (1.0) | 5.3 | 1.8 | o | n/a | n/a |

Data on maximum tolerated semaglutide dose (brackets indicate those still up-titrating dose), occurrence of adverse events and their clinical outcomes regarding delay in dose increase or dose decrease, in all participants (n=14). Participants were split into two groups, those who had ≤5% weight loss (n=12) or >5% weight loss (n=2) in the seven months prior to starting semaglutide treatment. Abbreviations: n/a, not applicable; o, no.

### 3.6. Glycaemic Control

Across *all* participants (including those without T2DM), there were no statistically significant changes in HbA1c either *prior* to (n=12, +2.9 ± 24.9 mmol/mol, +0.3 ± 2.3%, P=0.69, d +0.12) or *following* semaglutide initiation (n=11, -6.9 ± 33.4 mmol/mol, -0.6 ± 3.1%, P=0.51, d -0.21), and the difference between pre- and post-treatment periods was not significant (n=11, P=0.48, Cohen’s d -0.22) (**Table 3**, **Figure 3A**).

**Figure 3.**
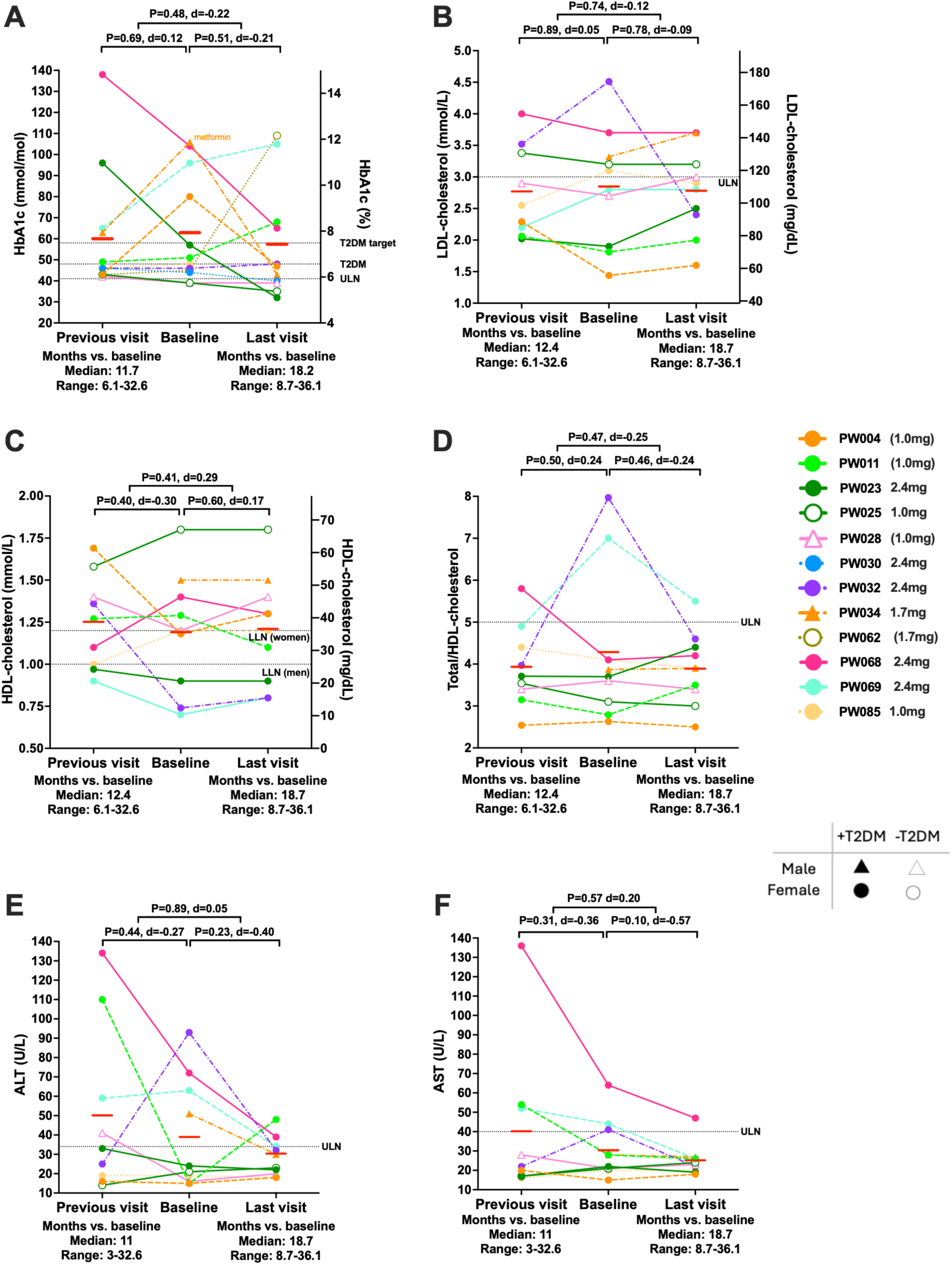
Clinical outcomes in adults with PWS treated with semaglutide. Graphs depict individual level data across three time points (visit previous to baseline (pre-treatment), baseline (treatment initiation), and last available follow-up), for (A) HbA1c, n=11; non-fasting (B) LDL-cholesterol, n=10; (C) HDL-cholesterol, n=10; (D) total/HDL-cholesterol ratio, n=10; (E) ALT, n=10; (F) AST, n=10, in those with ≤ 5% weight loss over seven months before starting semaglutide. Each graph displays individual participant trajectories, with semaglutide doses noted for each participant. Doses in brackets indicate participants still undergoing up-titration. Mean values are indicated by red lines. Median and range of time (in months) from baseline are given below x-axis for visits. Statistical analysis used repeated measured ANOVA with mixed model analysis using post-hoc Fisher LSD test, and paired t-test for comparison of change in outcome variable pre-visit vs. baseline and baseline vs. post-visit, with P-values and Cohen’s d effect sizes displayed: *P<0.05, **P<0.01. Abbreviations: ALT, alanine aminotransferase; ANOVA, repeated-measures analysis of variance; AST, aspartate aminotransferase; C, cholesterol; HbA1c, glycated haemoglobin; HDL, high-density lipoprotein; LDL, low-density lipoprotein; LLN, lower limit of normal; T2DM, type 2 diabetes mellitus; ULN, upper limit of normal.

**Figure 4.**
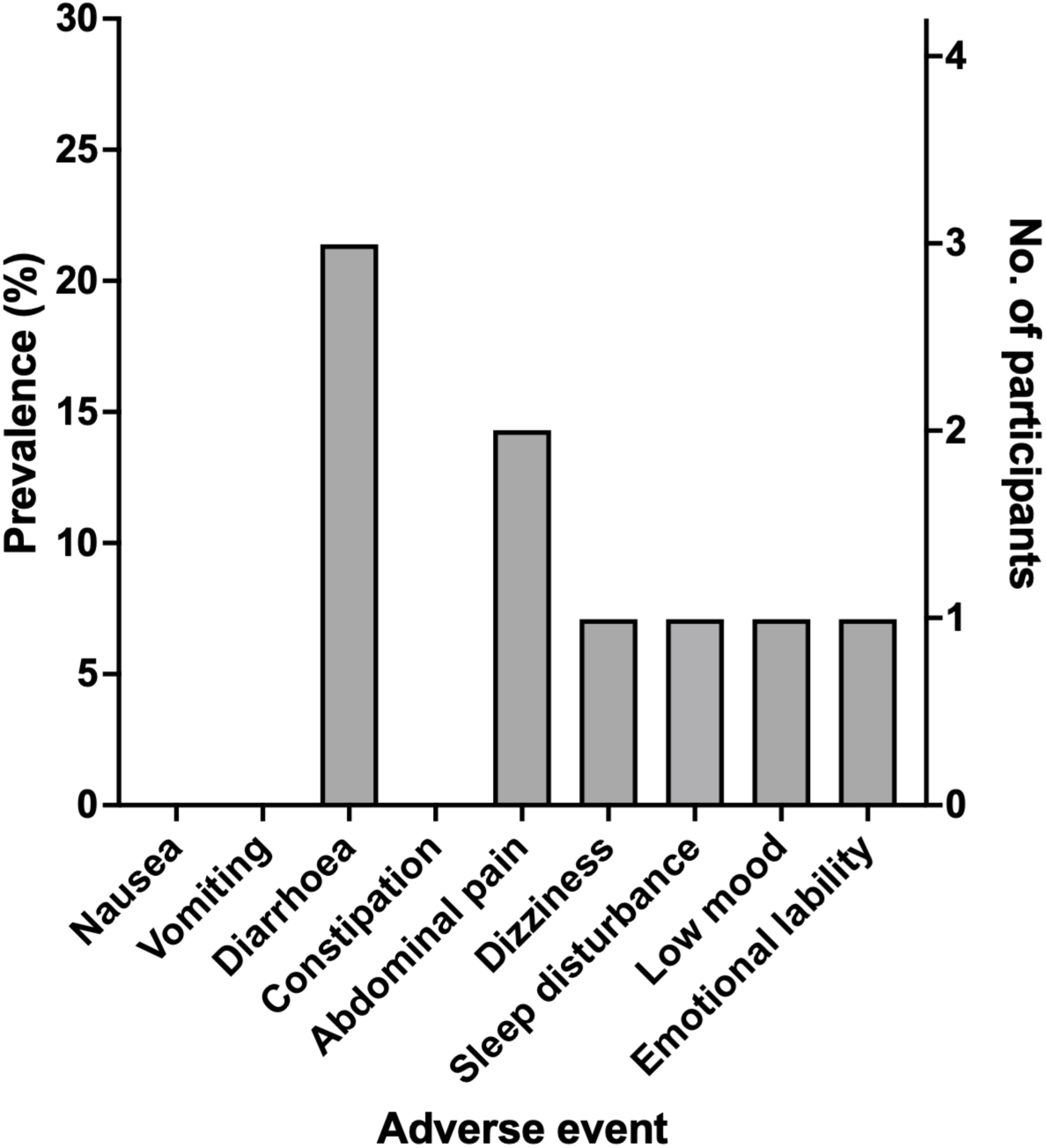
Adverse event outcomes in participants with PWS treated with semaglutide. Bar graph showing prevalence of adverse events following treatment with semaglutide in whole cohort (n=14).

However, among the five participants with *elevated* HbA1c at *baseline* (>48 mmol/mol, >6.5%) excluding the one participant who started metformin at baseline, three demonstrated reductions in HbA1c, with one achieving value within the normal range (**Figure 3A**). In these participants, there was no statistically significant change in HbA1c *prior* to semaglutide (n=5, -0.6 ± 35.4 mmol/mol, -0.1 ± 3.2%, P=0.97, d -0.02).

Although not reaching statistical significance, there was a moderate effect size for decrease in HbA1c *following* semaglutide treatment (n=5, -14.2 ± 25.5 mmol/mol, -1.3 ± 2.3%, P=0.28, d -0.56), and no statistically significant difference between pre- and post-treatment periods (n=5, P=0.43, d -0.39) (**Table 3**, **Figure 3A**).

### 3.7. Lipid Profile

No significant changes were observed in lipid parameters, including non-fasting LDL-cholesterol, HDL-cholesterol, or total/HDL-cholesterol ratio, either prior to or following semaglutide initiation, or between the pre- and post-treatment periods (**Table 3**, **Figure 3B-D**). However, very few had dyslipidaemia at baseline. Among the five participants with elevated LDL-cholesterol at baseline, three showed reductions after treatment with semaglutide, with two normalising (**Figure 3B**). Among the four participants with low HDL-cholesterol at baseline, three increased after treatment with semaglutide, with one normalising (**Figure 3C**). Both participants with elevated total/HDL-cholesterol at baseline had decreases after treatment with semaglutide, with one normalising (**Figure 3D**).

### 3.8. Liver Enzymes

Similarly, no statistically significant changes were observed in liver enzymes (ALT or AST) prior to or following treatment initiation, or between the pre- and post-treatment periods (**Table 3**, **Figure 3E-F**). However, baseline elevations in ALT and AST were uncommon. All four participants with elevated ALT at baseline had decreases after treatment with semaglutide, with three normalising (**Figure 3E**). All three participants with elevated AST at baseline, had decreases after treatment with semaglutide, with two normalising (**Figure 3F**).

## 4. DISCUSSION

This retrospective, observational cohort provides, as far as is known, the largest published data to date on the use of semaglutide in adults with PWS and is the first to report quantitative changes in hyperphagia using a validated questionnaire (HQ-CT). In 12 adults with PWS (nine with T2DM) treated with semaglutide, who had weight loss ≤5% in the 7 months prior to starting semaglutide, outcomes were generally as hypothesised, with:

i. overall duration of semaglutide treatment was median 17.2 months (range 8.7-36.1), with median semaglutide dose 2.4mg once weekly (1.0-2.4), with median time on highest dose 6.5 months (range 2.6-29.4), with 2/12 participants still up-titrating dose;
ii. 4/10 of patients reaching their highest dose did not tolerate the maximum dose 2.4mg weekly, but there were no discontinuations due to adverse effects;
iii. no significant weight loss on semaglutide compared to baseline, but a significant attenuation of pre-treatment weight gain, with similar results when examining weight change per month to account for variable follow-up durations;
iv. significant improvements in hyperphagia, as evident by reductions in mean HQ-CT score by 7.3/36, which had been stable prior to starting semaglutide (with similar changes in HQ-CT sub-scales of behaviour, drive, severity), without any change in the food environment as measured by the FSZ questionnaire;
v. improvements in HbA1c in the six participants with elevated HbA1c at baseline, decreasing in 4/6, and normalising in 1/4;
vi. no significant changes in serum LDL-cholesterol, HDL-cholesterol, total/HDL-cholesterol ratio, ALT or AST, but in those with abnormal levels at baseline, values usually improved, and often normalised;
vii. no serious side effects apart from 1/14 of all patients receiving treatment with semaglutide (including the two with weight loss prior to semaglutide) being diagnosed with ulcerative colitis, with an overall side effect profile, predominantly gastrointestinal, similar to that expected from non-PWS cohorts, though with no reported nausea, vomiting or constipation.

### 4.1. Body Weight

Prior to treatment initiation, participants were on average gaining weight, whereas weight remained stable on average following the introduction of semaglutide, though there was no significant weight loss on average. Even weight stabilisation is of benefit in PWS giving the natural history of progressive and often rapid weight gain and deterioration of obesity-related complications in adults with PWS (36).

These findings are broadly consistent with the limited observational literature on semaglutide use in PWS, although the magnitude of weight loss in our cohort appears more modest. Previously published reports of semaglutide in PWS described variability in changes of body weight, from +4.9% to −14.4% over treatment durations of 11–30 months (**Table 1**). For example, a single-case report of a man with obesity and T2DM treated with semaglutide 1 mg for 12 months demonstrated a 5.2% weight loss (103), while a small case series of four adults with PWS and T2DM reported a mean weight loss of 4.7% over a longer duration (25.5 months) at doses of 0.5–1 mg (104). Notably, greater weight reductions have been described in case reports of PWS without T2DM, including a series of three adults achieving a mean weight loss of 8.7% over 26 months with doses up to 2 mg (105). A further single case reported a 10.1% weight loss over 24 months in a man with T2DM, albeit at a relatively low dose (0.5 mg) (106). In comparison, the lower mean weight loss of 3.1% observed in our study on semaglutide, despite a longer mean treatment duration (19.6 months) and higher maximum dosing in some participants (up to 2.4 mg), may reflect differences in cohort composition, particularly the higher prevalence of T2DM (75%) and younger age. The predominance of females is unlikely to have contributed, as in non-PWS studies women lose more weight than men on GLP-1 analogues (114).

Furthermore, our findings align with prior observations suggesting a potentially attenuated weight loss response in individuals with T2DM (79). Notably, the stabilisation of weight following semaglutide initiation, in contrast to the marked upward trajectory observed prior to treatment, demonstrates the importance of considering pre-treatment weight trajectories when evaluating therapeutic response in observational studies in the absence of a placebo-control group.

### 4.2. Hyperphagia

The improvements in hyperphagia symptoms on semaglutide represent one of the most clinically relevant and novel findings of this study. This is an outcome not previously quantified in adults with PWS treated with semaglutide. This was assessed by the HQ-CT total score and was apparent across all three sub-domains of hyperphagic behaviour, drive and severity. However, HQ-CT scores are a subjective, self-reported assessment by carers over the previous two weeks, and can be influenced by reporting bias, natural variability, and regression to the mean. In the current open-label cohort study, the potential influence of the placebo effect must also be strongly acknowledged, since there may be pronounced placebo responses, particularly in domains related to behaviour, as seen in the DCCR trial, where HQ-CT scores improved in the placebo group (57).

A reduction in hyperphagia was also seen previously over 52 weeks in adolescents with PWS (though not adults) in a phase 2 placebo-controlled trial of the second generation GLP-1RA liraglutide, though no weight difference relative to placebo was seen (102). Hyperphagia is a defining and particularly burdensome feature of PWS, often leading to severe obesity and significant psychosocial challenges, with increased burden to caregivers and the family (21, 115).

Notably, HQ-CT scores remained relatively stable during the pre-treatment period, strengthening the likelihood that the observed changes were associated with treatment rather than natural fluctuation over time, though placebo effects cannot be excluded. Furthermore, FSZ scores did not change significantly during treatment with semaglutide in the subset with available data. As FSZ scores reflect the degree of environmental food restriction and supervision, the absence of change suggests that improvements in hyperphagia were unlikely to be attributable to alterations in the participants’ food environment. This finding supports a potential pharmacological effect of semaglutide on appetite regulation in individuals with PWS.

Although two participants had a change in social circumstances that may have affected weight and hyperphagia, with one switching schools (PW069), and the other moving residence from family home to supported living (PW032), the latter lost 11.3% weight over 30.6 months with HQ-CT decreasing by 10/36 points, despite their FSZ score falling from 67/80 to 58/80, indicating less strict control of food environment in supported living compared to the family home.

The average magnitude of reduction in HQ-CT total score (7.3/36 overall, 8.0/36 in those with HQ-CT >13/36 at baseline, 7.0/36 in those with baseline HQ-CT >22/36 at baseline) was comparable to that reported previously in trials of DCCR, a long-acting diazoxide preparation, that has now been licensed in USA for management of hyperphagia in PWS (57–60). In the initial placebo-controlled trial of DCCR in adolescents/adults with PWS, data prior to the interruption of the Covid-19 pandemic showed decreases in HQ-CT over 13 weeks of 6.6/36 in DCCR and 3.5/36 in placebo group, while in the whole study for those with HQ-CT >22/36 at baseline, HQ-CT decreased by 9.7/36 vs. 4.3/36 in DCCR and placebo groups respectively (57). In an open label extension of DCCR over 52 weeks, HQ-CT scores reduced overall by 9.9/36, and by an even greater amount (15.2/36) in those with severe hyperphagia (HQ-CT >22/36) at baseline (57). Furthermore, in a placebo-controlled trial of methionine aminopeptidase 2 (MetAP2) inhibitor, beloranib, in adolescents/adults with PWS, hyperphagia was significantly reduced with a mean HQ-CT reduction of 7.7/36, associated with weight loss, but unfortunately this product had to be subsequently withdrawn because of adverse events and deaths due to venous thrombosis (two fatal events of pulmonary embolism, two deep vein thrombotic events) (46).

### 4.3. Metabolic Profile

Although no significant changes were observed in glycaemic or lipid parameters overall, several participants with baseline abnormalities demonstrated improvements, including reductions in HbA1c, LDL-cholesterol, and liver enzymes, and increases in HDL-cholesterol. The absence of statistically significant changes likely reflects the small cohort size and the relatively limited number of participants with abnormal baseline values. Nonetheless, the direction of change observed in individuals with metabolic abnormalities is consistent with the established metabolic benefits of GLP-1RA in the general population.

Assessment of cardiovascular parameters is an important component of evaluating the safety and broader clinical impact of semaglutide in PWS, particularly given that GLP-1RA are associated with reductions in blood pressure but increases in resting heart rate (111). In our study, however, we were unable to reliably assess these outcomes. Blood pressure and pulse measurements were obtained in routine clinics, where patients with PWS are often anxious or excitable, potentially leading to unreliable readings. Furthermore, measurements were not standardised, as they were not consistently performed after a period of rest or repeated to obtain averaged values.

### 4.4. Safety Profile

Less than half of the participants experienced at least one adverse event, with diarrhoea being the most reported symptom. This is consistent with the known gastrointestinal side-effect profile of GLP-1RAs, although nausea and vomiting, frequently the most reported adverse event in larger clinical trials, was not seen in any participants (91–93).

Delayed gastric emptying is a commonly reported feature in children and adults with PWS (116, 117), with case reports of severe gastric dilatation and rupture in PWS, including children as young as 5 years which can be life-threatening (5, 25, 118–120). In a survey of 154 patients with PWS who had died, 3% were due to gastric rupture related to over-eating (119). In a large PWS cohort in the USA that died during 1973-2015, gastrointestinal related problems such as perforation, distension or obstruction were the causes of death in 10%, with equal distribution over the ages (25).

Decreased ability to vomit is commonly reported in PWS, both in children and adults (121). Although, the reasons for this are not completely known, diaphragmatic, abdominal, and intercostal muscle hypotonia may be contributory. Choking may occur frequently and has been reported as a cause of death in 8% of older children and adults, according to family collected data (119), with predisposing factors for choking hyperphagia, thick saliva, weakness of pharyngeal muscles and gastro-oesophageal reflux. More recently, videofluoroscopy in patients with PWS aged 5-35 years showed dysphagia with food residue in the pharynx and oesophageal stasis in almost all patients (122).

Since GLP-1 and its analogues including semaglutide can delay gastric emptying and cause gastroparesis (123–131), there are concerns that semaglutide may exacerbate or result in gastroparesis in PWS, perhaps especially in those with T2DM, and thus increase the risk of gastric necrosis or rupture. Furthermore, the high pain threshold and reduced vomiting in PWS may mask symptoms of gastroparesis.

For example, we have previously performed gastric emptying studies using Tc-99 semi-solid meal nuclear medicine scans to measure emptying half-life (t_1/2_) in seven adults with PWS and T2DM (**Supplementary Table S2**). Gastric emptying was markedly delayed in 1/6 patients not on liraglutide (t_1/2_ >180 min, normal <60 min), while the 7^th^ case already on liraglutide had mildly delayed gastric emptying t_1/2_ 79 mins, precluding further dose increase. Gastric emptying was slowed by liraglutide in 3 cases, becoming abnormal in two cases (16 to 348 min, 48 to 83 min, 25 to 49 min, normal <60 min).

Although the previous clinical trial of liraglutide in PWS did not result in any serious adverse events related to gastrointestinal issues, no formal gastric emptying assessments were performed (102). The lack of any symptoms suggestive of marked delay in gastric emptying in the current and previous observational case reports of semaglutide is reassuring, but given the greater efficacy of semaglutide over liraglutide, it will be important to consider risk of gastroparesis, including possible inclusion of formal assessment, during any future larger clinical trials of semaglutide (and other GLP-1RA) in PWS to confirm safety.

Interestingly, one participant had a new diagnosis of ulcerative colitis requiring dose reduction during treatment with semaglutide. There is a suggestion from observational cohorts in T2DM that the prevalence of ulcerative colitis might be higher in those on GLP-1RA, although this did not reach significance (132).

Among the six participants who reported adverse events, 16.6% required delayed dose escalation and 66.6% required a dose reduction due to side effects. Despite this, no participants discontinued semaglutide, which is notable given that discontinuation rates in the STEP trials for obesity typically ranged from 3–6% (**Supplementary Table S1**), and 20-60% in real world observational data (though often complicated by patients having to self-fund medication). This suggests that semaglutide was generally well tolerated in this cohort of adults with PWS, with the advised slow up-titration of dose, which indeed was slightly longer than minimum 4 weekly recommended dose increments. The time taken to reach the highest dose of 2.4mg was median 9.2 months (range 4.6 - 19.6 months), instead of 16 weeks with 4 weekly dose escalations.

### 4.5. Strengths and Limitations

This study has several notable strengths. It includes a larger cohort of patients from one centre with PWS treated with semaglutide than previously reported in the literature, and examines outcomes following administration of higher doses, with many reaching a maximum tolerated dose of 2.4mg. Importantly, it presents the first evidence of reduced hyperphagia with semaglutide using the HQ-CT. The inclusion of follow-up data from both before and after initiation of GLP-1RA therapy allows for a more robust evaluation of treatment effects over time. Additionally, by considering the FSZ questionnaire, it has been demonstrated that improvements in hyperphagia and weight were not attributable to changes in food security and environment.

Despite these strengths, several limitations should be acknowledged. The study was only a retrospective, observational cohort study, and lacked a placebo control group, and the overall sample size was small, limiting generalisability and the power to detect more subtle effects. There was marked variability in observational timepoints across patients both before and after starting treatment for all outcomes. This was addressed for weight change by calculating percentage weight change per month rather than absolute change alone; however, this approach may not eliminate the influence of differential follow-up duration as the rate of weight loss is not linear but attenuates over time. There was also variability in the maximum tolerated semaglutide dose (with two participants still up-titrating), and the duration of treatment overall and on highest dose. The latter was relatively short with median 6.5 months relative to overall median follow-up duration of 17.2 months, reflecting the gradual doe up-titration and limited patient review visits. The cohort was also heterogenous in PWS genotype, age, sex, severity of obesity and hyperphagia, prior weight trajectories, presence and degree of obesity-related comorbidities including T2DM, prior/current use of GH and sex steroids, use of concomitant anti-psychotic medication, use of other T2DM medications, and residential location, with sample size too small to allow most sub-group analyses. However, this cohort does represent real world data outside of a clinical trial, though responses to semaglutide will be heterogenous.

As the majority of patients included had T2DM, it is unknown whether the observed weight loss effects would be even greater in those without T2DM as has been seen across studies in the non-PWS population with overweight/obesity without vs. with T2DM (79), and suggested by the sub-analysis of the few without T2DM in the current study. Body weight was the outcome measure rather than body composition, and up to 20% of weight loss on GLP-1RA can be due to loss of muscle rather than fat tissue (133). The study was mainly in women with PWS, and in recent meta-analysis GLP-1RA appear more effective for weight loss in women than men without PWS (114). This study also only included adults with PWS, and greater effects of liraglutide on hyperphagia were seen in adolescents than adults with PWS in previous trial (102). Semaglutide has beneficial effects for weight loss in adolescents without PWS with a similar safety profile (74) (**Supplementary Table S1**).

Some HQ-CT questionnaires were completed retrospectively, which may introduce recall bias and affect the reliability of those responses, and accurate heart rate and blood pressure data was unavailable. HQ-CT questionnaires are also a self-reported outcome by carers and can be subject to reporter bias, natural variability, placebo effects, and changes in the food environment. Reassuringly in the sub-set with available data, the food environment appeared stable as assessed by the Food Safe Zone, but the influence of non-drug effects on hyperphagia cannot be excluded. Furthermore, four participants had potential confounds in their management which may have influenced outcomes, including two starting or changing doses of medications, and two moving residence or educational establishment.

### 4.6. Implications and Future Directions

Longer-term follow-up is needed to assess the durability of weight, and behavioural, cardiovascular and metabolic benefits, impact on quality of life, and potential for later dose reduction or maintenance strategies. Placebo-controlled trials of semaglutide in PWS, including individuals without T2DM, and adolescents and even younger children are a logical next step. Such trials should include measures of body composition, e.g. using dual-energy X-ray absorptiometry (DXA), hyperphagia using HQ-CT, behavioural and emotional problems using validated questionnaires (e.g. PWS-Profile), control of the food environment (FSZ), insulin resistance, glycaemic control, liver function tests, lipids, microalbuminuria, heart rate, blood pressure, and adverse events (111).

In the UK, recent NICE guidelines in 2026 have recommended early introduction of GLP-1 analogues in adults with early-onset T2DM, especially with overweight/obesity, not only for improvement in glycaemic control but because of cardiovascular and renal benefits (www.nice.org.uk/guidance/ng28). It may therefore not be ethical to assign adults with PWS and T2DM to placebo-treatment in clinical trials, as use of GLP-1 analogues should now be part of established therapy, though safety does still need to be confirmed.

Furthermore, the recent availability of higher dose semaglutide (7.2mg once weekly) in Europe and USA, which may produce additional weight loss over 2.4mg once weekly, supports consideration of using this higher dose in PWS (134, 135). Tirzepatide the dual GLP-1/GIP analogue appears more effective for weight loss than semaglutide in non-PWS overweight/obesity PWS (136). An open-label trial of tirzepatide in adults with PWS and obesity without uncontrolled T2DM is ongoing (https://clinicaltrials.gov/study/NCT06901245). Placebo-controlled studies of tirzepatide may be warranted based on their findings, together with collection of further real-world observational data given the increasing use of tirzepatide in the management of early-onset T2DM in PWS.

In addition, there is a large pipeline of additional GLP-1 and incretin-based therapies in development, but not yet licensed, which also appear more effective than semaglutide for weight loss in non-PWS cohorts with overweight/obesity, but have yet to be used in PWS, including the triple GLP-1/GIP/glucagon analogue, retatrutide, and combined GLP-1/amylin analogues, such as semaglutide/cagrilintide and amycretin (137–140).

### 4.7. Conclusion

This retrospective, observational cohort study provides the most comprehensive evaluation to date of semaglutide up to 2.4mg once weekly dose in adults with PWS, a population with complex obesity and limited treatment options. Semaglutide was associated with weight maintenance despite their prior weight gain, though not on average weight loss, reductions in hyperphagia, and improvements in glycaemic control, lipid profile, and liver enzymes. Importantly, semaglutide was well tolerated, with no discontinuations due to adverse effects, supporting its feasibility in routine care. Improvements in hyperphagia, supported by stable food security environments, suggest pharmacological efficacy beyond environmental or behavioural modification.

Nevertheless, limitations including small sample size, patient heterogeneity, open-label nature with lack of a placebo-control group, and variability in duration of follow-up, time on highest dose, and achieved dose, warrants cautious interpretation. Larger, prospective, longer-term, placebo-controlled trials with standardised dose titration periods and follow-up duration, are urgently needed to confirm efficacy, assess durability of response, and evaluate behavioural outcomes and obesity-related phenotypes, in adolescents and adults with PWS, especially in those without T2DM. The pipeline of combination incretin and other gut hormone-based therapies for obesity may produce pharmacological agents with even greater efficacy than semaglutide.

## DATA AVAILABILITY STATEMENT

Datasets generated and/or analysed during the current study are available from the corresponding author on reasonable request.

## REFERENCES

1. Goldstone AP, Holland AJ, Hauffa BP, Hokken-Koelega AC, Tauber M. Recommendations for the diagnosis and management of Prader-Willi syndrome. J Clin Endocrinol Metab. (2008) 93:4183–97.

2. Miller JL, Driscoll DC, Lynn CH, Goldstone AP, Kimonis V, Dykens E, Butler MG, Shuster JJ, Driscoll DJ. Nutritional phases in Prader-Willi syndrome. Am J Med Genet A. (2011) 155:1040–9.

3. Butler MG, Kimonis V, Dykens E, Gold JA, Miller J, Tamura R, Driscoll DJ. Prader-Willi syndrome and early-onset morbid obesity NIH rare disease consortium: A review of natural history study. Am J Med Genet A. (2018) 176:368–75.

4. Manzardo AM, Loker J, Heinemann J, Loker C, Butler MG. Survival trends from the Prader-Willi Syndrome Association (USA) 40-year mortality survey. Genet Med. (2018) 20:24–30.

5. Bellis SA, Kuhn I, Adams S, Mullarkey L, Holland A. The consequences of hyperphagia in people with Prader-Willi Syndrome: A systematic review of studies of morbidity and mortality. Eur J Med Genet. (2022) 65:104379.

6. Shaikh MG, Barrett TG, Bridges N, Chung R, Gevers EF, Goldstone AP, Holland A, Kanumakala S, Krone R, Kyriakou A, Livesey EA, Lucas-Herald AK, Meade C, Passmore S, Roche E, Smith C, Soni S. Prader-Willi syndrome: guidance for children and transition into adulthood. Endocr Connect. (2024) 13:e240091.

7. Driscoll DJ, Miller JL, Cassidy SB. Prader-Willi Syndrome. In: Adam MP, Feldman J, Mirzaa GM, Pagon RA, Wallace SE, Bean LJH, et al., editors. Gene Reviews. Seattle, WA (1993).

8. Goldstone AP. Prader-Willi syndrome: advances in genetics, pathophysiology and treatment. Trends Endocrinol Metab. (2004) 15:12–20.

9. Manning KE, Holland AJ. Puzzle Pieces: Neural Structure and Function in Prader-Willi Syndrome. Diseases. (2015) 3:382–415.

10. Azor AM, Cole JH, Holland AJ, Dumba M, Patel MC, Sadlon A, Goldstone AP, Manning KE. Increased brain age in adults with Prader-Willi syndrome. Neuroimage Clin. (2019) 21:101664.

11. Tauber M, Hoybye C. Endocrine disorders in Prader-Willi syndrome: a model to understand and treat hypothalamic dysfunction. Lancet Diabetes Endocrinol. (2021) 9:235–46.

12. Bochukova EG. Transcriptomics of the Prader-Willi syndrome hypothalamus. Handb Clin Neurol. (2021) 181:369–79.

13. Brown SSG, Manning KE, Fletcher P, Holland A. In vivo neuroimaging evidence of hypothalamic alteration in Prader-Willi syndrome. Brain Commun. (2022) 4:fcac229.

14. Butler JV, Whittington JE, Holland AJ, McAllister CJ, Goldstone AP. The transition between the phenotypes of Prader-Willi syndrome during infancy and early childhood. Dev Med Child Neurol. (2010) 52:e88–93.

15. Whittington J, Holland A. A review of psychiatric conceptions of mental and behavioural disorders in Prader-Willi syndrome. Neurosci Biobehav Rev. (2018) 95:396–405.

16. Schwartz L, Caixas A, Dimitropoulos A, Dykens E, Duis J, Einfeld S, Gallagher L, Holland A, Rice L, Roof E, Salehi P, Strong T, Taylor B, Woodcock K. Behavioral features in Prader-Willi syndrome (PWS): consensus paper from the International PWS Clinical Trial Consortium. J Neurodev Disord. (2021) 13:25.

17. Dykens EM, Maxwell MA, Pantino E, Kossler R, Roof E. Assessment of hyperphagia in Prader-Willi syndrome. Obesity. (2007) 15:1816–26.

18. Meade C, Martin R, McCrann A, Lyons J, Meehan J, Hoey H, Roche E. Prader-Willi Syndrome in children: Quality of life and caregiver burden. Acta Paediatr. (2021) 110:1665–70.

19. Bos-Roubos A, Wingbermuhle E, Biert A, de Graaff L, Egger J. Family Matters: Trauma and Quality of Life in Family Members of Individuals With Prader-Willi Syndrome. Front Psychiatry. (2022) 13:897138.

20. Howell TA, Matza LS, Mallya UG, Goldstone AP, Butsch WS, Lazarus E. Health state utilities associated with hyperphagia: Data for use in cost-utility models. Obes Sci Pract. (2023) 9:376–82.

21. Mazaheri MM, Rae-Seebach RD, Preston HE, Schmidt M, Kountz-Edwards S, Field N, Cassidy S, Packman W. The impact of Prader-Willi syndrome on the family’s quality of life and caregiving, and the unaffected siblings’ psychosocial adjustment. J Intellect Disabil Res. (2013) 57:861–73.

22. Caliandro P, Grugni G, Padua L, Kodra Y, Tonali P, Gargantini L, Ragusa L, Crino A, Taruscio D. Quality of life assessment in a sample of patients affected by Prader-Willi syndrome. J Paediatr Child Health. (2007) 43:826–30.

23. Kayadjanian N, Schwartz L, Farrar E, Comtois KA, Strong TV. High levels of caregiver burden in Prader-Willi syndrome. PLoS One. (2018) 13:e0194655.

24. Whittington JE, Holland AJ, Webb T. Ageing in people with Prader-Willi syndrome: mortality in the UK population cohort and morbidity in an older sample of adults. Psychological medicine. (2015) 45:615–21.

25. Butler MG, Manzardo AM, Heinemann J, Loker C, Loker J. Causes of death in Prader-Willi syndrome: Prader-Willi Syndrome Association (USA) 40-year mortality survey. Genet Med. (2017) 19:635–42.

26. Butler MG, Miller JL, Forster JL. Prader-Willi Syndrome - Clinical Genetics, Diagnosis and Treatment Approaches: An Update. Curr Pediatr Rev. (2019) 15:207–44.

27. Wolfe G, Salehi V, Browne A, Riddle R, Hall E, Fam J, Tichansky D, Myers S. Metabolic and bariatric surgery for obesity in Prader Willi syndrome: systematic review and meta-analysis. Surg Obes Relat Dis. (2023) 19:907–15.

28. Sinnema M, Maaskant MA, van Schrojenstein Lantman-de Valk HM, van Nieuwpoort IC, Drent ML, Curfs LM, Schrander-Stumpel CT. Physical health problems in adults with Prader-Willi syndrome. Am J Med Genet A. (2011) 155A:2112–24.

29. O’Neill LP, Murray LE. Anxiety and depression symptomatology in adult siblings of individuals with different developmental disability diagnoses. Res Dev Disabil. (2016) 51–52:116-25.

30. Tan Q, Orsso CE, Deehan EC, Triador L, Field CJ, Tun HM, Han JC, Muller TD, Haqq AM. Current and emerging therapies for managing hyperphagia and obesity in Prader-Willi syndrome: A narrative review. Obes Rev. (2020) 21:e12992.

31. Grugni G, Sartorio A, Crino A, Fintini D. The pharmacological management of obesity in Prader-Willi syndrome: what does the future hold? Expert Opin Pharmacother. (2026) 27:389–92.

32. Miller JL, Tan M. Dietary Management for Adolescents with Prader-Willi Syndrome. Adolesc Health Med Ther. (2020) 11:113–8.

33. Alsaif M, Elliot SA, MacKenzie ML, Prado CM, Field CJ, Haqq AM. Energy Metabolism Profile in Individuals with Prader-Willi Syndrome and Implications for Clinical Management: A Systematic Review. Adv Nutr. (2017) 8:905–15.

34. Bellicha A, Coupaye M, Mosbah H, Tauber M, Oppert JM, Poitou C. Physical Activity in Patients with Prader-Willi Syndrome-A Systematic Review of Observational and Interventional Studies. J Clin Med. (2021) 10:2528.

35. Grugni G, Crino A, Bosio L, Corrias A, Cuttini M, De TT, Di BE, Franzese A, Gargantini L, Greggio N, Iughetti L, Livieri C, Naselli A, Pagano C, Pozzan G, Ragusa L, Salvatoni A, Trifiro G, Beccaria L, Bellizzi M, Bellone J, Brunani A, Cappa M, Caselli G, Cerioni V, Delvecchio M, Giardino D, Ianni F, Memo L, Pilotta A, Pomara C, Radetti G, Sacco M, Sanzari A, Sartorio A, Tonini G, Vettor R, Zaglia F, Chiumello G. The Italian National Survey for Prader-Willi syndrome: an epidemiologic study. Am J Med Genet A. (2008) 146:861–72.

36. Vrana-Diaz CJ, Balasubramanian P, Kayadjanian N, Bohonowych J, Strong TV. Variability and change over time of weight and BMI among adolescents and adults with Prader-Willi syndrome: a 6-month text-based observational study. Orphanet J Rare Dis. (2020) 15:233.

37. Pellikaan K, Rosenberg AGW, Kattentidt-Mouravieva AA, Kersseboom R, Bos-Roubos AG, Veen-Roelofs JMC, van Wieringen N, Hoekstra FME, van den Berg SAA, van der Lely AJ, de Graaff LCG. Missed Diagnoses and Health Problems in Adults With Prader-Willi Syndrome: Recommendations for Screening and Treatment. J Clin Endocrinol Metab. (2020) 105:e4671–87.

38. Morales JS, Valenzuela PL, Pareja-Galeano H, Rincon-Castanedo C, Rubin DA, Lucia A. Physical exercise and Prader-Willi syndrome: A systematic review. Clin Endocrinol (Oxf). (2019) 90:649–61.

39. Hughes BM, Holland A, Hodebeck-Stuntebeck N, Garrick L, Goldstone AP, Lister M, Moore C, Hughes M. Body weight, behaviours of concern, and social contact in adults and adolescents with Prader-Willi syndrome in full-time care services: Findings from pooled international archival data. Orphanet J Rare Dis. (2024) 19:48.

40. Hirsch HJ, Benarroch F, Genstil L, Pollak Y, Derei D, Forer D, Mastey Ben-Yehuda H, Gross-Tsur V. Long-term weight control in adults with Prader-Willi syndrome living in residential hostels. Am J Med Genet A. (2021) 185:1175–81.

41. Frixou M, Vlek D, Lucas-Herald AK, Keir L, Kyriakou A, Shaikh MG. The use of growth hormone therapy in adults with Prader-Willi syndrome: A systematic review. Clin Endocrinol (Oxf). (2021) 94:645–55.

42. Lucas-Herald AK, Perry CG, Shaikh MG. Review of growth hormone therapy in adolescents and young adults with Prader-Willi syndrome. Expert review of endocrinology & metabolism. (2015) 10:259–67.

43. Miller JL, Linville TD, Dykens EM. Effects of metformin in children and adolescents with Prader-Willi syndrome and early-onset morbid obesity: a pilot study. J Pediatr Endocrinol Metab. (2014) 27:23–9.

44. Mahmoud R, Kimonis V, Butler MG. Clinical Trials in Prader-Willi Syndrome: A Review. Int J Mol Sci. (2023) 24:2150.

45. Motaghedi R, Lipman EG, Hogg JE, Christos PJ, Vogiatzi MG, Angulo MA. Psychiatric adverse effects of rimonobant in adults with Prader Willi syndrome. Eur J Med Genet. (2011) 54:14–8.

46. McCandless SE, Yanovski JA, Miller J, Fu C, Bird LM, Salehi P, Chan CL, Stafford D, Abuzzahab MJ, Viskochil D, Barlow SE, Angulo M, Myers SE, Whitman BY, Styne D, Roof E, Dykens EM, Scheimann AO, Malloy J, Zhuang D, Taylor K, Hughes TE, Kim DD, Butler MG. Effects of MetAP2 inhibition on hyperphagia and body weight in Prader-Willi syndrome: A randomized, double-blind, placebo-controlled trial. Diabetes Obes Metab. (2017) 19:1751–61.

47. Allas S, Caixas A, Poitou C, Coupaye M, Thuilleaux D, Lorenzini F, Diene G, Crino A, Illouz F, Grugni G, Potvin D, Bocchini S, Delale T, Abribat T, Tauber M. AZP-531, an unacylated ghrelin analog, improves food-related behavior in patients with Prader-Willi syndrome: A randomized placebo-controlled trial. PLoS One. (2018) 13:e0190849.

48. Millendo Therapeutics Inc. Effects of Livoletide (AZP-531) on Food-related Behaviors in Patients With Prader-Willi Syndrome (ZEPHYR). (2021). https://clinicaltrials.gov/study/NCT03790865.

49. Miller JL, Lacroix A, Bird LM, Shoemaker AH, Haqq A, Deal CL, Clark KA, Ames MH, Suico JG, de la Pena A, Fortier C. The Efficacy, Safety, and Pharmacology of a Ghrelin O-Acyltransferase Inhibitor for the Treatment of Prader-Willi Syndrome. J Clin Endocrinol Metab. (2022) 107:e2373–e80.

50. Rhythm Pharmaceuticals Inc. A Ph 2, Randomized, Double-Blind, Placebo-Controlled Pilot Study to Assess the Effects of RM-493, a Melanocortin 4 Receptor (MC4R) Agonist, in Obese Subjects with Prader-Willi Syndrome (PWS) on Safety, Weight Reduction, and Food-Related Behaviors. (2023). https://clinicaltrialsgov/ct2/show/NCT02311673.

51. Einfeld SL, Smith E, McGregor IS, Steinbeck K, Taffe J, Rice LJ, Horstead SK, Rogers N, Hodge MA, Guastella AJ. A double-blind randomized controlled trial of oxytocin nasal spray in Prader Willi syndrome. Am J Med Genet A. (2014) 164A:2232–9.

52. Kuppens RJ, Donze SH, Hokken-Koelega AC. Promising effects of oxytocin on social and food-related behaviour in young children with Prader-Willi syndrome: a randomized, double-blind, controlled crossover trial. Clin Endocrinol (Oxf). (2016) 85:979–87.

53. Damen L, Grootjen LN, Juriaans AF, Donze SH, Huisman TM, Visser JA, Delhanty PJD, Hokken-Koelega ACS. Oxytocin in young children with Prader-Willi syndrome: Results of a randomized, double-blind, placebo-controlled, crossover trial investigating 3 months of oxytocin. Clin Endocrinol (Oxf). (2021) 94:774–85.

54. Dykens EM, Miller J, Angulo M, Roof E, Reidy M, Hatoum HT, Willey R, Bolton G, Korner P. Intranasal carbetocin reduces hyperphagia in individuals with Prader-Willi syndrome. JCI Insight. (2018) 3:e98333.

55. Roof E, Deal CL, McCandless SE, Cowan RL, Miller JL, Hamilton JK, Roeder ER, McCormack SE, Roshan Lal TR, Abdul-Latif HD, Haqq AM, Obrynba KS, Torchen LC, Vidmar AP, Viskochil DH, Chanoine JP, Lam CKL, Pierce MJ, Williams LL, Bird LM, Butler MG, Jensen DE, Myers SE, Oatman OJ, Baskaran C, Chalmers LJ, Fu C, Alos N, McLean SD, Shah A, Whitman BY, Blumenstein BA, Leonard SF, Ernest JP, Cormier JW, Cotter SP, Ryman DC. Intranasal Carbetocin Reduces Hyperphagia, Anxiousness, and Distress in Prader-Willi Syndrome: CARE-PWS Phase 3 Trial. J Clin Endocrinol Metab. (2023) 108:1696–708.

56. Matesevac L, Vrana-Diaz CJ, Bohonowych JE, Schwartz L, Strong TV. Analysis of Hyperphagia Questionnaire for Clinical Trials (HQ-CT) scores in typically developing individuals and those with Prader-Willi syndrome. Sci Rep. (2023) 13:20573.

57. Miller JL, Gevers E, Bridges N, Yanovski JA, Salehi P, Obrynba KS, Felner EI, Bird LM, Shoemaker AH, Angulo M, Butler MG, Stevenson D, Abuzzahab J, Barrett T, Lah M, Littlejohn E, Mathew V, Cowen NM, Bhatnagar A, Investigators DP. Diazoxide Choline Extended-Release Tablet in People With Prader-Willi Syndrome: A Double-Blind, Placebo-Controlled Trial. J Clin Endocrinol Metab. (2023) 108:1676–85.

58. Strong TV, Miller JL, McCandless SE, Gevers E, Yanovski JA, Matesevac L, Bohonowych J, Ballal S, Yen K, Hirano P, Cowen NM, Bhatnagar A. Behavioral changes in patients with Prader-Willi syndrome receiving diazoxide choline extended-release tablets compared to the PATH for PWS natural history study. J Neurodev Disord. (2024) 16:22.

59. Miller JL, Gevers E, Bridges N, Yanovski JA, Salehi P, Obrynba KS, Felner EI, Bird LM, Shoemaker AH, Angulo M, Butler MG, Stevenson D, Goldstone AP, Wilding J, Lah M, Shaikh MG, Littlejohn E, Abuzzahab MJ, Fleischman A, Hirano P, Yen K, Cowen NM, Bhatnagar A, Investigators CC. Diazoxide choline extended-release tablet in people with Prader-Willi syndrome: results from long-term open-label study. Obesity (Silver Spring). (2024) 32:252–61.

60. Miller JL, Bridges N, Felner EI, Salehi P, Yanovski JA, Stevenson DA, Mejia-Corletto J, Shaikh MG, Abuzzahab J, Fleischman A, Kimonis V, Shoemaker AH, Holland A, Bird LM, Obrynba KS, Lah M, Littlejohn E, Harwood K, Shea H, Viskochil D, Hirano P, Yen K, Ballal S, Huang M, Cowen NM, Bhatnagar A, Gevers E. Diazoxide Choline Extended-Release Tablets in Prader-Willi Syndrome: A Randomized, Double-Blind, Withdrawal Period Study. J Clin Endocrinol Metab. (2026) doi: 10.1210/clinem/dgaf661.

61. Scheimann AO, Butler MG, Gourash L, Cuffari C, Klish W. Critical analysis of bariatric procedures in Prader-Willi syndrome. J Pediatr Gastroenterol Nutr. (2008) 46:80–3.

62. Kjems LL, Holst JJ, Volund A, Madsbad S. The influence of GLP-1 on glucose-stimulated insulin secretion: effects on beta-cell sensitivity in type 2 and nondiabetic subjects. Diabetes. (2003) 52:380–6.

63. Liu QK. Mechanisms of action and therapeutic applications of GLP-1 and dual GIP/GLP-1 receptor agonists. Front Endocrinol. (2024) 15:1431292.

64. Maselli DB, Camilleri M. Effects of GLP-1 and Its Analogs on Gastric Physiology in Diabetes Mellitus and Obesity. Adv Exp Med Biol. (2021) 1307:171–92.

65. Astrup A. Reflections on the discovery GLP-1 as a satiety hormone: Implications for obesity therapy and future directions. Eur J Clin Nutr. (2024) 78:551–6.

66. Drucker DJ. Efficacy and Safety of GLP-1 Medicines for Type 2 Diabetes and Obesity. Diabetes Care. (2024) 47:1873–88.

67. Engel JA, Jerlhag E. Role of appetite-regulating peptides in the pathophysiology of addiction: implications for pharmacotherapy. CNS Drugs. (2014) 28:875–86.

68. Holst JJ. GLP-1 physiology in obesity and development of incretin-based drugs for chronic weight management. Nat Metab. (2024) 6:1866–85.

69. Tamayo-Trujillo R, Ruiz-Pozo VA, Cadena-Ullauri S, Guevara-Ramirez P, Paz-Cruz E, Zambrano-Villacres R, Simancas-Racines D, Zambrano AK. Molecular mechanisms of semaglutide and liraglutide as a therapeutic option for obesity. Front Nutr. (2024) 11:1398059.

70. Johansen VBI, Petersen J, Lund J, Mathiesen CV, Fenselau H, Clemmensen C. Brain control of energy homeostasis: Implications for anti-obesity pharmacotherapy. Cell. (2025) 188:4178–212.

71. Wilding JPH, Batterham RL, Calanna S, Davies M, Van Gaal LF, Lingvay I, McGowan BM, Rosenstock J, Tran MTD, Wadden TA, Wharton S, Yokote K, Zeuthen N, Kushner RF, Group SS. Once-Weekly Semaglutide in Adults with Overweight or Obesity. N Engl J Med. (2021) 384:989–1002.

72. Wadden TA, Bailey TS, Billings LK, Davies M, Frias JP, Koroleva A, Lingvay I, O’Neil PM, Rubino DM, Skovgaard D, Wallenstein SOR, Garvey WT, Investigators S. Effect of Subcutaneous Semaglutide vs Placebo as an Adjunct to Intensive Behavioral Therapy on Body Weight in Adults With Overweight or Obesity: The STEP 3 Randomized Clinical Trial. JAMA. (2021) 325:1403–13.

73. Rubino D, Abrahamsson N, Davies M, Hesse D, Greenway FL, Jensen C, Lingvay I, Mosenzon O, Rosenstock J, Rubio MA, Rudofsky G, Tadayon S, Wadden TA, Dicker D, Investigators S. Effect of Continued Weekly Subcutaneous Semaglutide vs Placebo on Weight Loss Maintenance in Adults With Overweight or Obesity: The STEP 4 Randomized Clinical Trial. JAMA. (2021) 325:1414–25.

74. Weghuber D, Barrett T, Barrientos-Perez M, Gies I, Hesse D, Jeppesen OK, Kelly AS, Mastrandrea LD, Sorrig R, Arslanian S, Investigators ST. Once-Weekly Semaglutide in Adolescents with Obesity. N Engl J Med. (2022) 387:2245–57.

75. Garvey WT, Batterham RL, Bhatta M, Buscemi S, Christensen LN, Frias JP, Jodar E, Kandler K, Rigas G, Wadden TA, Wharton S, Group SS. Two-year effects of semaglutide in adults with overweight or obesity: the STEP 5 trial. Nat Med. (2022) 28:2083–91.

76. Kadowaki T, Isendahl J, Khalid U, Lee SY, Nishida T, Ogawa W, Tobe K, Yamauchi T, Lim S, investigators S. Semaglutide once a week in adults with overweight or obesity, with or without type 2 diabetes in an east Asian population (STEP 6): a randomised, double-blind, double-dummy, placebo-controlled, phase 3a trial. Lancet Diabetes Endocrinol. (2022) 10:193–206.

77. Rubino DM, Greenway FL, Khalid U, O’Neil PM, Rosenstock J, Sorrig R, Wadden TA, Wizert A, Garvey WT, Investigators S. Effect of Weekly Subcutaneous Semaglutide vs Daily Liraglutide on Body Weight in Adults With Overweight or Obesity Without Diabetes: The STEP 8 Randomized Clinical Trial. JAMA. (2022) 327:138–50.

78. Davies M, Faerch L, Jeppesen OK, Pakseresht A, Pedersen SD, Perreault L, Rosenstock J, Shimomura I, Viljoen A, Wadden TA, Lingvay I, Group SS. Semaglutide 2.4 mg once a week in adults with overweight or obesity, and type 2 diabetes (STEP 2): a randomised, double-blind, double-dummy, placebo-controlled, phase 3 trial. Lancet. (2021) 397:971–84.

79. Lingvay I, Agarwal S. A revolution in obesity treatment. Nat Med. (2023) 29:2406–8.

80. Gibbons C, Blundell J, Tetens Hoff S, Dahl K, Bauer R, Baekdal T. Effects of oral semaglutide on energy intake, food preference, appetite, control of eating and body weight in subjects with type 2 diabetes. Diabetes Obes Metab. (2021) 23:581–8.

81. Wharton S, Batterham RL, Bhatta M, Buscemi S, Christensen LN, Frias JP, Jodar E, Kandler K, Rigas G, Wadden TA, Garvey WT. Two-year effect of semaglutide 2.4 mg on control of eating in adults with overweight/obesity: STEP 5. Obesity. (2023) 31:703–15.

82. Kolotkin RL, Jeppesen OK, Baker-Knight J, Lee SY, Tokita A, Kadowaki T. Effect of once-weekly subcutaneous semaglutide 2.4 mg on weight- and health-related quality of life in an East Asian population: Patient-reported outcomes from the STEP 6 trial. Clin Obes. (2023) 13:e12589.

83. Rubino D, Bjorner JB, Rathor N, Sharma AM, von Huth Smith L, Wharton S, Wadden T, Zeuthen N, Kolotkin RL. Effect of semaglutide 2.4 mg on physical functioning and weight- and health-related quality of life in adults with overweight or obesity: Patient-reported outcomes from the STEP 1-4 trials. Diabetes Obes Metab. (2024) 26:2945–55.

84. Robert SA, Rohana AG, Shah SA, Chinna K, Wan Mohamud WN, Kamaruddin NA. Improvement in binge eating in non-diabetic obese individuals after 3 months of treatment with liraglutide - A pilot study. Obes Res Clin Pract. (2015) 9:301–4.

85. Chao AM, Wadden TA, Walsh OA, Gruber KA, Alamuddin N, Berkowitz RI, Tronieri JS. Effects of Liraglutide and Behavioral Weight Loss on Food Cravings, Eating Behaviors, and Eating Disorder Psychopathology. Obesity (Silver Spring). (2019) 27:2005–10.

86. Da Porto A, Casarsa V, Colussi G, Catena C, Cavarape A, Sechi L. Dulaglutide reduces binge episodes in type 2 diabetic patients with binge eating disorder: A pilot study. Diabetes Metab Syndr. (2020) 14:289–92.

87. Richards J, Bang N, Ratliff EL, Paszkowiak MA, Khorgami Z, Khalsa SS, Simmons WK. Successful treatment of binge eating disorder with the GLP-1 agonist semaglutide: A retrospective cohort study. Obes Pillars. (2023) 7:100080.

88. Aoun L, Almardini S, Saliba F, Haddadin F, Mourad O, Jdaidani J, Morcos Z, Al Saidi I, Bou Sanayeh E, Saliba S, Almardini M, Zaidan J. GLP-1 receptor agonists: A novel pharmacotherapy for binge eating (Binge eating disorder and bulimia nervosa)? A systematic review. J Clin Transl Endocrinol. (2024) 35:100333.

89. Allison KC, Chao AM, Bruzas MB, McCuen-Wurst C, Jones E, McAllister C, Gruber K, Berkowitz RI, Wadden TA, Tronieri JS. A pilot randomized controlled trial of liraglutide 3.0 mg for binge eating disorder. Obes Sci Pract. (2023) 9:127–36.

90. White RT, Henriquez P, Innocent B, Bullers K, Elmaoued A. Incretin-Based Therapies for the Treatment of Binge Eating - A Systematic Review. Pharmacotherapy. (2026) 46:e70135.

91. Jalleh RJ, Rayner CK, Hausken T, Jones KL, Camilleri M, Horowitz M. Gastrointestinal effects of GLP-1 receptor agonists: mechanisms, management, and future directions. Lancet Gastroenterol Hepatol. (2024) 9:957–64.

92. Ismaiel A, Scarlata GGM, Boitos I, Leucuta DC, Popa SL, Al Srouji N, Abenavoli L, Dumitrascu DL. Gastrointestinal adverse events associated with GLP-1 RA in non-diabetic patients with overweight or obesity: a systematic review and network meta-analysis. Int J Obes (Lond). (2025) 49:1946–57.

93. Wharton S, Calanna S, Davies M, Dicker D, Goldman B, Lingvay I, Mosenzon O, Rubino DM, Thomsen M, Wadden TA, Pedersen SD. Gastrointestinal tolerability of once-weekly semaglutide 2.4 mg in adults with overweight or obesity, and the relationship between gastrointestinal adverse events and weight loss. Diabetes Obes Metab. (2022) 24:94–105.

94. Van Laren J, Friesleben C, Gershovich O, Helm L, Patel RJ, Delate T. Characterisation of real-world patients who discontinued a glucagon-like peptide-1 agonist. Diabetes Obes Metab. (2026) 28:3965–73.

95. Drucker DJ. The benefits of GLP-1 drugs beyond obesity. Science. (2024) 385:258–60.

96. Lu Y, Chen J, Guo Y, Ding H, Liu YL, Van Name MA, Sharifi M, Lu Y, Chen Y. Cardiometabolic Profiles of Oral and Subcutaneous Glucagon-Like Peptide-1 Receptor Mono-Agonists in Adults With Overweight or Obesity: A Systematic Review and Network Meta-Analysis. Diabetes Obes Metab. (2026) 10.1111/dom.70742.

97. Rico-Fontalvo J, Daza-Arnedo R, Elbert A, Correa-Rotter R, Dina-Batlle E, Lorca-Herrera E, de Moraes TP, Sanchez-Polo V, Builes-Montano CE. Renal Outcomes of GLP-1 Receptor Agonists and Tirzepatide Across CKD Stages and Metabolic Phenotypes (Type 2 Diabetes and/or Overweight/Obesity): A Scoping Review. Diabetes Ther. (2026) 17:499–528.

98. Sanyal AJ, Newsome PN, Kliers I, Ostergaard LH, Long MT, Kjaer MS, Cali AMG, Bugianesi E, Rinella ME, Roden M, Ratziu V, Group ES. Phase 3 Trial of Semaglutide in Metabolic Dysfunction-Associated Steatohepatitis. N Engl J Med. (2025) 392:2089–99.

99. Kim YM, Lee YJ, Kim SY, Cheon CK, Lim HH. Successful rapid weight reduction and the use of liraglutide for morbid obesity in adolescent Prader-Willi syndrome. Ann Pediatr Endocrinol Metab. (2020) 25:52–6.

100. Cyganek K, Koblik T, Kozek E, Wojcik M, Starzyk J, Malecki MT. Liraglutide therapy in Prader-Willi syndrome. Diabetic medicine: a journal of the British Diabetic Association. (2011) 28:755–6.

101. Senda M, Ogawa S, Nako K, Okamura M, Sakamoto T, Ito S. The glucagon-like peptide-1 analog liraglutide suppresses ghrelin and controls diabetes in a patient with Prader-Willi syndrome. Endocr J. (2012) 59:889–94.

102. Diene G, Angulo M, Hale PM, Jepsen CH, Hofman PL, Hokken-Koelega A, Ramesh C, Turan S, Tauber M. Liraglutide for Weight Management in Children and Adolescents with Prader-Willi Syndrome and Obesity. J Clin Endocrinol Metab. (2022) 108:4–12.

103. Sani E, Prato GD, Zenti MG, Bordugo A, Trombetta M, Bonora E. Effects of Semaglutide on Glycemic Control and Weight Loss in a Patient with Prader-Willi Syndrome: A Case Report. Endocr Metab Immune Disord Drug Targets. (2022) 22:1053–7.

104. Gimenez-Palop O, Romero A, Casamitjana L, Pareja R, Rigla M, Caixas A. Effect of semaglutide on weight loss and glycaemic control in patients with Prader-Willi Syndrome and type 2 diabetes. Endocrinol Diabetes Nutr. (2024) 71:83–7.

105. Koceva A, Mlekus Kozamernik K, Janez A, Herman R, Ferjan S, Jensterle M. Case report: Long-term efficacy and safety of semaglutide in the treatment of syndromic obesity in Prader Willi syndrome - case series and literature review. Front Endocrinol (Lausanne). (2024) 15:1528457.

106. Dinoi E, Daniele G, Michelucci A, Baldinotti F, Campi F, Marchetti P, Del Prato S, Dardano A. Efficacy and safety of once-weekly semaglutide monotherapy in a young subject with Prader-Willi syndrome, obesity, and type 2 diabetes: a case report. Front Endocrinol (Lausanne). (2025) 16:1533209.

107. Gokul PR, Apperley L, Parkinson J, Clark K, Lund K, Owens M, Senniappan S. Semaglutide, a Long-Acting GLP-1 Analogue, for the Management of Early-Onset Obesity due to MC4R Defect: A Case Report. Horm Res Paediatr. (2025) 98:148–55.

108. Bhatnagar P, Ahmad NN, Li X, Coghlan M, Kaplan LM, Farooqi IS. Tirzepatide leads to weight reduction in people with obesity due to MC4R deficiency. Nat Med. (2025) 31:3294–6.

109. Ferch M, Peitsch I, Kautzky-Willer A, Greber-Platzer S, Stattermayer AF, Krebs M, Scherer T. Effectiveness of the Dual GIP/GLP1-Agonist Tirzepatide in 2 Cases of Alstrom Syndrome, a Rare Obesity Syndrome. J Clin Endocrinol Metab. (2025) 110:3364–9.

110. Salama M, Hassan D, Vairo FPE, Lteif A, Hentz R, Olson O, Pittock S, Al Nofal A, Creo A, Kumar S. Effects of Semaglutide on BMI and Cardiometabolic Profile in Adolescents With Variants in Monogenic Obesity-Related Genes. Pediatr Obes. (2026) 21:e70091.

111. Finer N, Strong T, Vrana-Diaz C, Stafford DEJ. Recommendations for real-world evidence of efficacy and safety of GLP-1 agonists in Prader-Willi syndrome: Report of a workshop held by the Foundation for Prader-Willi Research and International Prader Willi Syndrome Organisation. Diabetes Obes Metab. (2026) 28:799–802.

112. Sterling RK, Lissen E, Clumeck N, Sola R, Correa MC, Montaner J, M SS, Torriani FJ, Dieterich DT, Thomas DL, Messinger D, Nelson M, Investigators AC. Development of a simple noninvasive index to predict significant fibrosis in patients with HIV/HCV coinfection. Hepatology. (2006) 43:1317–25.

113. Dykens EM, Roof E, Hunt-Hawkins H, Strong TV. Validation of the Food Safe Zone questionnaire for families of individuals with Prader-Willi syndrome. J Neurodev Disord. (2025) 17:6.

114. Alexander GC, Xiao X, Dilek S, Lewis S, Deng Q, Kim M, Bolanle D, Saldanha IJ, Mehta HB. Heterogeneity of Treatment Effects of Glucagon-Like Peptide-1 Receptor Agonists for Weight Loss in Adults: A Systematic Review and Meta-Analysis. JAMA Intern Med. (2026):e258222.

115. Howell MJ, Schenck CH, Crow SJ. A review of nighttime eating disorders. Sleep medicine reviews. (2009) 13:23–34.

116. Hoybye C, Barkeling B, Naslund E, Thoren M, Hellstrom PM. Eating behavior and gastric emptying in adults with Prader-Willi syndrome. Ann Nutr Metab. (2007) 51:264–9.

117. Arenz T, Schwarzer A, Pfluger T, Koletzko S, Schmidt H. Delayed gastric emptying in patients with Prader Willi Syndrome. J Pediatr Endocrinol Metab. (2010) 23:867–71.

118. Wharton RH, Wang T, GraemeCook F, Briggs S, Cole RE. Acute idiopathic gastric dilatation with gastric necrosis in individuals with Prader-Willi syndrome. Am J Med Genet. (1997) 73:437–41.

119. Stevenson DA, Heinemann J, Angulo M, Butler MG, Loker J, Rupe N, Kendell P, Cassidy SB, Scheimann A. Gastric rupture and necrosis in Prader-Willi syndrome. J Pediatr Gastroenterol Nutr. (2007) 45:272–4.

120. Blat C, Busquets E, Gili T, Caixas A, Gabau E, Corripio R. Gastric Dilatation and Abdominal Compartment Syndrome in a Child with Prader-Willi Syndrome. Am J Case Rep. (2017) 18:637–40.

121. Holm VA, Cassidy SB, Butler MG, Hanchett JM, Greenswag LR, Whitman BY, Greenberg F. Prader-Willi syndrome: consensus diagnostic criteria. Pediatrics. (1993) 91:398–402.

122. Gross RD, Gisser R, Cherpes G, Hartman K, Maheshwary R. Subclinical dysphagia in persons with Prader-Willi syndrome. Am J Med Genet A. (2017) 173:384–94.

123. Naslund E, Gutniak M, Skogar S, Rossner S, Hellstrom PM. Glucagon-like peptide 1 increases the period of postprandial satiety and slows gastric emptying in obese men. AmJClinNutr. (1998) 68:525–30.

124. Naslund E, Bogefors J, Skogar S, Gryback P, Jacobsson H, Holst JJ, Hellstrom PM. GLP-1 slows solid gastric emptying and inhibits insulin, glucagon, and PYY release in humans. American Journal of Physiology. (1999) 277:R910–R6.

125. Flint A, Raben A, Ersboll AK, Holst JJ, Astrup A. The effect of physiological levels of glucagon-like peptide-1 on appetite, gastric emptying, energy and substrate metabolism in obesity. Int J Obes Relat Metab Disord. (2001) 25:781–92.

126. DeFronzo RA, Okerson T, Viswanathan P, Guan X, Holcombe JH, MacConell L. Effects of exenatide versus sitagliptin on postprandial glucose, insulin and glucagon secretion, gastric emptying, and caloric intake: a randomized, cross-over study. Curr Med Res Opin. (2008) 24:2943–52.

127. van Can J, Sloth B, Jensen CB, Flint A, Blaak EE, Saris WH. Effects of the once-daily GLP-1 analog liraglutide on gastric emptying, glycemic parameters, appetite and energy metabolism in obese, non- diabetic adults. Int J Obes (Lond). (2014) 38:784–93.

128. Modi R, Rye P, Cawsey S, Birch DW, Sharma AM. Liraglutide Effects on Upper Gastrointestinal Investigations: Implications Prior to Bariatric Surgery. Obes Surg. (2018) 28:2113–6.

129. Hjerpsted JB, Flint A, Brooks A, Axelsen MB, Kvist T, Blundell J. Semaglutide improves postprandial glucose and lipid metabolism, and delays first-hour gastric emptying in subjects with obesity. Diabetes Obes Metab. (2018) 20:610–9.

130. Sherwin M, Hamburger J, Katz D, DeMaria S, Jr. Influence of semaglutide use on the presence of residual gastric solids on gastric ultrasound: a prospective observational study in volunteers without obesity recently started on semaglutide. Can J Anaesth. (2023) 70:1300–6.

131. Camilleri M, Carlson P, Dilmaghani S. Prevalence and variations in gastric emptying delay in response to GLP-1 receptor agonist liraglutide. Obesity (Silver Spring). (2024) 32:232–3.

132. Xie Y, Choi T, Al-Aly Z. Mapping the effectiveness and risks of GLP-1 receptor agonists. Nat Med. (2025) 31:951–62.

133. Anyiam O, Ardavani A, Rashid RSA, Panesar A, Idris I. How do glucagon-like Peptide-1 receptor agonists affect measures of muscle mass in individuals with, and without, type 2 diabetes: A systematic review and meta-analysis. Obes Rev. (2025) 26:e13916.

134. Lingvay I, Bergenheim SJ, Buse JB, Freitas P, Garvey WT, Harder-Lauridsen NM, Rosenstock J, Sahu K, Wharton S, group SUTDt. Once-weekly semaglutide 7.2 mg in adults with obesity and type 2 diabetes (STEP UP T2D): a randomised, controlled, phase 3b trial. Lancet Diabetes Endocrinol. (2025) 13:935–48.

135. Wharton S, Freitas P, Hjelmesaeth J, Kabisch M, Kandler K, Lingvay I, Quiroga M, Rosenstock J, Garvey WT, group SUt. Once-weekly semaglutide 7.2 mg in adults with obesity (STEP UP): a randomised, controlled, phase 3b trial. Lancet Diabetes Endocrinol. (2025) 13:949–63.

136. Aronne LJ, Horn DB, le Roux CW, Ho W, Falcon BL, Gomez Valderas E, Das S, Lee CJ, Glass LC, Senyucel C, Dunn JP, Investigators S-T. Tirzepatide as Compared with Semaglutide for the Treatment of Obesity. N Engl J Med. (2025) 393:26–36.

137. Jastreboff AM, Kaplan LM, Frias JP, Wu Q, Du Y, Gurbuz S, Coskun T, Haupt A, Milicevic Z, Hartman ML, Retatrutide Phase 2 Obesity Trial I. Triple-Hormone-Receptor Agonist Retatrutide for Obesity - A Phase 2 Trial. N Engl J Med. (2023) 389:514–26.

138. Garvey WT, Bluher M, Osorto Contreras CK, Davies MJ, Winning Lehmann E, Pietilainen KH, Rubino D, Sbraccia P, Wadden T, Zeuthen N, Wilding JPH, Group RS. Coadministered Cagrilintide and Semaglutide in Adults with Overweight or Obesity. N Engl J Med. (2025) 393:635–47.

139. Davies MJ, Bajaj HS, Broholm C, Eliasen A, Garvey WT, le Roux CW, Lingvay I, Lyndgaard CB, Rosenstock J, Pedersen SD, Group RS. Cagrilintide-Semaglutide in Adults with Overweight or Obesity and Type 2 Diabetes. N Engl J Med. (2025) 393:648–59.

140. Dahl K, Toubro S, Dey S, Duque do Vale R, Flint A, Gasiorek A, Heydorn A, Jastreboff AM, Key C, Petersen SB, Vegge A, Adelborg K. Amycretin, a novel, unimolecular GLP-1 and amylin receptor agonist administered subcutaneously: results from a phase 1b/2a randomised controlled study. Lancet. (2025) 406:149–62.

