## Supplementary Material for "Efficacy and safety of semaglutide for obesity and hyperphagia in adults with Prader-Willi syndrome"

Supplementary Table 1. Summary of STEP clinical trials of semaglutide for overweight/obesity.

Supplementary Table 2. Tc-99 gastric emptying findings in adults with PWS and type 2 diabetes mellitus.

Questions from the Hyperphagia Questionnaire for Clinical Trials (HQ-CT)

| Trial | Author | Year | Population BMI status | Treatment or withdrawal | T2DM | Age group | % T2DM | Other Rx | Group | N start | N end | Dropout % | Age (y), mean ± SD | Maximum dose (mg) | Duration (weeks) | Start weight | BMI baseline (kg/m²) | Δ mean weight (kg) | mean diff kg vs. placebo | P | Δ mean % weight | mean diff % wt loss vs. placebo | P | % ≥5% weight loss | % ≥10% weight loss | % ≥15% weight loss | % ≥20% weight loss |
|---|---|---|---|---|---|---|---|---|---|---|---|---|---|---|---|---|---|---|---|---|---|---|---|---|---|---|---|
| STEP 1 | Wilding | 2021 | OW/OB | treatment | o | adult | 0 | lifestyle | Semaglutide | 1306 | 1059 | 18.9 | 46 ± 13 | 2.4 | 68 | 105.4 ± 22.1 | 37.8 ± 6.7 | -15.3 | -12.7 | <0.001 | -14.9 | -12.5 | <0.001 | 86.4 | 69.1 | 50.5 | 32 |
| | | | | | | | | | Placebo | 655 | 499 | 23.8 | 47 ±12 | | | 105.2 ± 21.5 | 38.0 ± 6.5 | -2.6 | | | -2.4 | | | 31.5 | 12 | 4.9 | 1.7 |
| STEP 2 | Davies | 2021 | OW/OB | treatment | yes | adult | 100 | lifestyle | Semaglutide | 404 | 387 | 4.2 | >18 | 2.4 | 68 | 99.9 ± 22.5 | 35.9 ± 6.4 | -9.7 | -6.2 | | -9.6 | -6.2 | <0.0001 | 68.8 | 45.6 | 25.8 | |
| | | | | | | | | | Placebo | 403 | 375 | 6.9 | >18 | | | 100.5 ± 20.9 | 35.9 ± 6.5 | -3.5 | | | -3.4 | | | 28.5 | 28.7 | 13.7 | |
| STEP 3 | Wadden | 2021 | OW/OB | treatment | o | adult | 0 | LCD 8w | Semaglutide | 407 | 373 | 8.4 | 46 ± 13 | 2.4 | 68 | 106.9 ± 22.8 | 38.1 ± 6.7 | -16.8 | -10.6 | <0.001 | -16 | -10.3 | <0.001 | 86.6 | 75.3 | 55.8 | 35.7 |
| | | | | | | | | plus behav therapy | Placebo | 204 | 189 | 7.4 | 46 ± 13 | | | 103.7 ± 22.9 | 37.8 ± 6.9 | -6.2 | | | -5.7 | | | 47.6 | 27 | 13.2 | 3.7 |
| STEP 4 | Rubino | 2021 | OW/OB | withdrawal | o | adult | 0 | lifestyle | Semaglutide | 535 | 518 | 3.2 | 47 ±12 | 2.4 | 48 | 96.5 ± 22.5 | 34.5 ± 6.9 | -7.1 | -13.2 | <0.001 | -7.9 | -14.8 | <0.001 | | | | |
| | | | | | | | | | Placebo | 268 | 248 | 7.5 | 46 ± 12 | | | 95.4 ± 22.7 | 34.1 ± 7.1 | 6.1 | | | 6.9 | | | | | | |
| STEP 5 | Garvey | 2022 | OW/OB | treatment 2y | o | adult | 0 | lifestyle | Semaglutide | 152 | 144 | 5.3 | 47 ±12 | 2.4 | 104 | 105.6 ± 20.8 | 38.6 ± 6.7 | -16.1 | -12.9 | | -15.2 | -12.6 | <0.0001 | 77.1 | 61.8 | 52.1 | 36.1 |
| | | | | | | | (42-51% preDM) | | Placebo | 152 | 128 | 15.8 | 47 ±10 | | | 106.5 ± 23.1 | 38.5 ± 7.2 | -3.2 | | | -2.6 | | | 34.4 | 13.3 | 7 | 2.3 |
| STEP 6 | Kadowaki | 2022 | OW/OB | treatment | some | adult | 25 | lifestyle | Semaglutide | 199 | 193 | 3.0 | 52 ± 12 | 2.4 | 68 | 86.9 ± 16.5 | 32.0 ± 4.6 | | | | -13.2 | -11.1 | <0.0001 | 82.9 | 60.6 | 40.9 | |
| | | | | | | (East Asian) | | | Placebo (pooled) | 101 | 100 | 1.0 | 50 ± 9 | | | 90.2 ± 15.1 | 31.9 ± 4.2 | | | | -2.1 | | | 21 | 41.8 | 24.5 | |
| STEP 8 | Rubino | 2022 | OW/OB | treatment | o | adult | 0 | lifestyle | Semaglutide | 126 | 117 | 7.1 | 48 ±14 | 2.4 | 68 | 102.5 ± 25.3 | 37.0 ± 7.4 | -15.3 | -13.7 | | -15.8 | -13.9 | | 87.2 | 70.9 | 55.6 | 38.5 |
| | | | | | | | | | Placebo | 85 | 78 | 8.2 | 51 ± 12 | | | 103.7 ± 22.5 | 37.2 ± 6.4 | -1.6 | | | -1.9 | | | 29.5 | 15.4 | 6.4 | 2.6 |
| STEPTEENS | Weghuber | 2022 | OB | treatment | few | adolescent | 4 | lifestyle | Semaglutide | 134 | 131 | 2.2 | 15.5 ± 1.5 | 2.4 | 68 | 109.9 ± 25.2 | 37.7 ± 6.7 | -5.8 | -5.9 | | -16.1 | -16.7 | <0.001 | 72.5 | 61.8 | 53.4 | 37.4 |
| | | | | | | 12-18y | | | Placebo | 67 | 62 | 7.5 | 15.3 ± 1.6 | | | 102.6 ± 22.3 | 35.7 ± 5.4 | 0.1 | | | 0.6 | | | 17.7 | 8.1 | 4.8 | 3.2 |

**Supplementary Table 1. Summary of STEP clinical trials of semaglutide for overweight/obesity.**

Abbreviations: BMI, body mass index; o, no; OB, obesity; OW, overweight; preDM, prediabetes; Rx, prescription; SD, standard deviation; T2DM, type 2 diabetes mellitus; y, years. Data is taken from the following clinical trials:


Garvey WT, Batterham RL, Bhatta M, Buscemi S, Christensen LN, Frias JP, Jodar E, Kandler K, Rigas G, Wadden TA, Wharton S, Group SS. Two-year effects of semaglutide in adults with overweight or obesity: the STEP 5 trial. Nat Med. 28:2083-2091, 2022.

Kadowaki T, Isendahl J, Khalid U, Lee SY, Nishida T, Ogawa W, Tobe K, Yamauchi T, Lim S, investigators S. Semaglutide once a week in adults with overweight or obesity, with or without type 2 diabetes in an east Asian population (STEP 6): a randomised, double-blind, double-dummy, placebo-controlled, phase 3a trial. Lancet Diabetes Endocrinol. 10:193-206, 2022.

Rubino D, Abrahamsson N, Davies M, Hesse D, Greenway FL, Jensen C, Lingvay I, Mosenzon O, Rosenstock J, Rubio MA, Rudofsky G, Tadayon S, Wadden TA, Dicker D, Investigators S. Effect of Continued Weekly Subcutaneous Semaglutide vs Placebo on Weight Loss Maintenance in Adults With Overweight or Obesity: The STEP 4 Randomized Clinical Trial. JAMA. 325:1414-1425, 2021.

Rubino DM, Greenway FL, Khalid U, O'Neil PM, Rosenstock J, Sorrig R, Wadden TA, Wizert A, Garvey WT, Investigators S. Effect of Weekly Subcutaneous Semaglutide vs Daily Liraglutide on Body Weight in Adults With Overweight or Obesity Without Diabetes: The STEP 8 Randomized Clinical Trial. JAMA. 327:138-150, 2022.

Wadden TA, Bailey TS, Billings LK, Davies M, Frias JP, Koroleva A, Lingvay I, O'Neil PM, Rubino DM, Skovgaard D, Wallenstein SOR, Garvey WT, Investigators S. Effect of Subcutaneous Semaglutide vs Placebo as an Adjunct to Intensive Behavioral Therapy on Body Weight in Adults With Overweight or Obesity: The STEP 3 Randomized Clinical Trial. JAMA. 325:1403-1413, 2021.

Weghuber D, Barrett T, Barrientos-Perez M, Gies I, Hesse D, Jeppesen OK, Kelly AS, Mastrandrea LD, Sorrig R, Arslanian S, Investigators ST. Once-Weekly Semaglutide in Adolescents with Obesity. N Engl J Med. 387:2245-2257, 2022.

Wilding JPH, Batterham RL, Calanna S, Davies M, Van Gaal LF, Lingvay I, McGowan BM, Rosenstock J, Tran MTD, Wadden TA, Wharton S, Yokote K, Zeuthen N, Kushner RF, Group SS. Once-Weekly Semaglutide in Adults with Overweight or Obesity. N Engl J Med. 384:989-1002, 2021.

| Sex | Age range (yr) | BMI (kg/m2) | HbA1c (%, mmol/mol) | T2DM Rx | Gastric emptying t1/2 baseline (NR <60 mins) | Gastric emptying t1/2 on Liraglutide (NR <60 mins) | Liraglutide dose (mg) | Clinical decision about GLP-1 analogue |
|---|---|---|---|---|---|---|---|---|
| F | 18-22 | 39.5 | 9.6%, 81 | Met Th I | 48 | 83 | 1.8 | Continue GLP-1RA |
| F | 18-22 | 30.7 | 6.8%, 51 | Met | 25 | 49 | 1.8 | Continue GLP-1RA |
| F | 43-47 | 41.4 | 13.3%, 122 | Met, I | - | 79 | 1.2 | GLP-1RA dose increase CI, started Th |
| M | 23-27 | 36.6 | 107 | Met, Th | 16 | 348 | 1.8 | GLP-IRA discontinued |
| M | 28-32 | 54 | 10.2%, 88 | ? | >180 | - | - | GLP-1RA CI |
| M | 23-27 | 40.4 | 9.9%, 85 | Met, Th | 32 | - | - | GLP-1RA not CI |
| M | 18-22 | 64.3 | FBG 4.9 mmol/L | Met, S | 16 | - | - | GLP-1RA not CI |

**Supplementary Table 2. Tc-99 gastric emptying findings in adults with PWS and type 2 diabetes mellitus.**

Abbreviations: BMI, body mass index; CI, contraindicated; F, female; FBG, fasting blood glucose; GLP-1RA, GLP-1 receptor agonist; I, Insulin; M, male; Met, metformin; mins, minutes; NR, normal range; Rx, treatment; S, sulphonylurea; T2DM, type 2 diabetes mellitus; Th, Thiazolidinedione; t1/2, half-life; yr, year.

Adapted from: Ali SN, Bridges N, Al-Nahhas A, Papadopoulou D, Sampson M, Goldstone AP. Assessing gastric emptying before and after Introduction of GLP-1 agonists for glycaemic and body weight control in Prader-Willi Syndrome. Abstracts 31st Annual Meeting of the Obesity Society, Atlanta USA, 2013.

**Questions from the Hyperphagia Questionnaire for Clinical Trials (HQ-CT)**

Instructions: The following items refer to the person in your care and assessment of his/her food-related behaviour during the past 2 weeks.

(1) During the past 2 weeks, how upset did the person generally become when denied a desired food?
___ Not at all upset
___ A little upset
___ Moderately upset
___ Very upset
___ Extremely upset

(2) During the past 2 weeks, how often did the person try to bargain or manipulate to get more food at meals?
___ Never
___ Up to 2 times a week
___ 3 to 6 times a week
___ Every day
___ Several times a day

(3) During the past 2 weeks, how often did the person forage through trash for food?
___ Never
___ 1 time
___ 2 times
___ 3 times
___ 4 or more times

(4) During the past 2 weeks, how often did the person get up at night to food seek?
___ Never
___ 1 time
___ 2 times
___ 3 times
___ 4 or more times

(5) During the past 2 weeks, how persistent was the person in asking or looking for food after being told “no” or “no more”?
___ Not at all persistent
___ A little persistent
___ Moderately persistent
___ Very persistent
___ Extremely persistent

(6) During the past 2 weeks, outside of normal meal times, how much time did the person generally spend asking or talking about food?
___ Less than 5 minutes a day
___ 5 to 15 minutes a day
___ 15 to 30 minutes a day
___ 30 minutes to 1 hour a day
___ More than 1 hour a day

(7) During the past 2 weeks, how often did the person try to sneak or steal food (that you are aware of)?
___ Never
___ 1 time
___ 2 times
___ 3 times
___ 4 or more times

(8) During the past 2 weeks, when others tried to stop the person from asking about food, how distressed did he or she generally appear?
___ Not at all distressed
___ A little distressed
___ Moderately distressed
___ Very distressed
___ Extremely distressed

(9) During the past 2 weeks, how often did food-related behavior interfere with the person's normal daily activities, such as self-care, recreation, school, or work?
___ Never
___ Up to 2 times a week
___ 3 to 6 times a week
___ Every day
___ Several times a day

Each question is scored 0-4.
HQ-CT total: questions 1-9 (score 0-36)
HQ-CT behaviour: questions 2, 3, 4 and 7 (score 0-16)
HQ-CT drive: questions 1, 5 and 8 (score 0-12)
HQ-CT severity: questions 6 and 9 (score 0-8)

Taken from: Matesevac L, Vrana-Diaz CJ, Bohonowych JE, Schwartz L, Strong TV. Analysis of Hyperphagia Questionnaire for Clinical Trials (HQ-CT) scores in typically developing individuals and those with Prader-Willi syndrome. *Sci Rep*. 13:20573, 2023.

Contact information for permission to use the HQ-CT: Foundation for Prader-Willi Research (www.fpwr.org),.
